# Effect of Acute Cardiovascular Exercise on Cerebral Blood Flow: A Systematic Review

**DOI:** 10.1101/2022.08.14.22278765

**Authors:** Lisa Mulser, David Moreau

## Abstract

A single bout of cardiovascular exercise can have a cascade of physiological effects, including increased blood flow to the brain. This effect has been documented across multiple modalities, yet studies have reported mixed findings. Here, we systematically review evidence for the acute effect of cardiovascular exercise on cerebral blood flow across a range of neuroimaging techniques and exercise characteristics. Based on 52 studies and a combined sample size of 1,174 individuals, our results indicate that the acute effect of cardiovascular exercise on cerebral blood flow generally follows an inverted U-shaped relationship, whereby blood flow increases early on but eventually decreases as exercise continues. However, we also find that this effect is not uniform across studies, instead varying across a number of key variables including exercise characteristics, brain regions, and neuroimaging modalities. As the most comprehensive synthesis on the topic to date, this systematic review sheds light on the determinants of exercise-induced change in cerebral blood flow, a necessary step toward personalized interventions targeting brain health across a range of populations.

## 1. Introduction

The human brain roughly comprises 2% of total body mass (Burma et al., 2020), yet it consumes up to 15% of cardiac output and 20% of total oxygen intake at rest (Williams & Leggett, 1989). Given the brain’s limited substrate storage (Ashley et al., 2020; Burma et al., 2020), maintaining nutrient and oxygen supply to cerebral tissue via continuous blood flow is key to brain health and cognition. Cerebral blood flow (CBF)—the volume of arterial blood (mL) transported to 100g of cerebral tissue per minute (Buxton, 2005)—provides a measure of how well these supplies are delivered to neural tissue. CBF, and thus brain tissue oxygenation (Andrianopoulos et al., 2018), is actively maintained by cerebral vasculature either dilating or contracting to facilitate nutrient and oxygen delivery (Willie et al., 2014). Constant blood supply to the brain is paramount—significant reductions in cerebral perfusion can lead to loss of consciousness (Smith et al., 2011; Van Lieshout et al., 2003), brain damage, or even death (Braz et al., 2017; Del Zoppo et al., 2011; Lipton, 1999; Willie et al., 2014).

### 1.1. Structural Overview of Cerebral Blood Flow

A series of anatomical structures enables sustained CBF. Sitting at the base of the brain at the junction of the carotid arteries, which supply blood from the heart, the circle of Willis plays a critical role (Thomas et al., 2020). Internal carotid arteries carry approximately 70 % of total CBF to the brainstem, the cortex, and the cerebellum, while the vertebral arteries supply the rest (Willie et al., 2014). Within neural tissue, internal carotid arteries further branch into cerebral arteries, which are key to maintaining cerebral perfusion (Willie et al., 2014). As part of this interrelated network of structures providing blood flow, the brain has a built-in mechanism to regulate and maintain quasi constant cerebral perfusion across varying levels of systemic blood pressure (Clausen et al., 2016; Lassen, 1959; Ogoh et al., 2005). This mechanism—cerebral autoregulation (CA)—shields the brain from drastic changes in mean arterial pressure, sympathetic nerve activity, and arterial CO_2_ levels (Clausen et al., 2016), thus enabling adequate neural function and cognitive performance at all times.

### 1.2. Major Factors Influencing Cerebral Blood Flow

Despite built-in regulatory mechanisms, CBF still varies significantly within and across individuals. Several attributes can be associated with these differences; for example, CBF is elevated during childhood to meet the developing brain’s increased neuronal and metabolic demands, but later declines in adulthood (Biagi et al., 2007). Throughout healthy adulthood, decreases in white matter, hormonal changes in the brain, and loss of cortical neurons and brain mass contribute to a reduction in CBF (Smirl et al., 2016). Toward later adulthood, the drop in CBF appears to mirror cognitive decline (Ellis et al., 2017).

CBF also differs between sexes (Satterthwaite et al., 2014; Tontisirin et al., 2007). For instance, Tontisirin and colleagues (2007) reported sex differences in regional perfusion patterns, with girls (4-8 years of age) having significantly higher middle cerebral artery flow velocity and basilar artery flow velocity than age-matched boys. However, these differences do not seem to affect overall CA, for which no sex differences were observed (Tontisirin et al., 2007). Satterthwaite and colleagues (2014) further note that global brain perfusion levels diverge significantly between sexes during puberty, suggesting that significant differences may only arise during that time.

A myriad of additional genetic and environmental factors can influence CBF, including smoking, lifestyle and dietary habits, and sleep patterns. Among these, cardiovascular exercise is unique because it can affect CBF via a wide range of mechanisms while also being associated with numerous health benefits. Exercise has been shown to slow the age-related decline in CBF (Ainslie et al., 2008; Bailey et al., 2013; Barnes, 2015; Colcombe & Kramer, 2003; Colcombe et al., 2006; Lucas et al., 2012), and cardiorespiratory fitness correlates with global CBF (gCBF) across the lifespan, with fit individuals showing higher gCBF than their age-matched untrained counterparts (Braz et al., 2017).

### 1.3. Acute Cardiovascular Exercise and Cerebral Blood Flow

Beyond the well-documented relationship between cardiovascular fitness, chronic exercise, and CBF (Anazodo et al., 2016; Chapman et al., 2013), a growing trend of literature has focused on the potent acute effect of cardiovascular exercise. Acute effects refer to transient outcomes that do not last beyond a few hours; these are typically measured after a single bout of exercise (Moreau & Chou, 2019). Cardiovascular exercise is the most effective type of regimen to elicit change in CBF, as it leads to increased blood flow across the entire body. Various exercise modalities have been proposed, most typically running, swimming, cycling, or rowing (Perentis et al., 2021). In studies focusing on acute effects, cardiovascular exercise typically ranges in intensity, from moderate (i.e., aerobic regimens) to intense (i.e., high-intensity regimens). Bouts of aerobic exercise are usually longer than high-intensity regimens and do not typically elicit oxygen deficits during the session. In contrast, high-intensity bouts are typically shorted and often interleaved with rest periods for recovery, as oxygen deficit is maximal and the body relies primarily on anaerobic metabolism (Klein et al., 2019; Labrecque et al., 2020).

A single bout of cardiovascular exercise affects CBF in a number of ways. One of the principal physiological mechanisms associated with exercise is an increase in heart rate to meet oxygen demands. An increased heart rate results in a higher blood flow that allows for enhanced oxygen, excitatory neurotransmitter, and nutrient supply to the brain (Olivo et al., 2021). This physiological response is thought to lead to cognitive enhancement, for example in the form of sharper attention or cognitive control (Olivo et al., 2021). Besides the need of the body and brain to meet nutrient demands in the face of cardiovascular exercise, Hiura and colleagues (2009) and Billinger and colleagues (2021) offer a different explanation for why exercise may increase CBF. These authors suggest that exercise may lead to an increase in cerebral perfusion due to the mental effort required and the associated increase in cortical activation (Billinger et al., 2021; Hiura et al., 2009). For instance, when a person exercises, they may consciously or subconsciously think of some of the movements they would like to engage in. In this context, motor simulation may add to the need for increased nutrient supply to the brain when exercising, due to the added energy requirements. Regardless of whether the mental effort associated with exercise drives the increase in CBF linked to cardiovascular exercise, Ogoh and Ainslie (2009) argue that changes in cerebral vascular tone (i.e., the degree to which a blood vessel constricts relative to its maximally dilated state) play an important role. Specifically, Ogoh and Ainslie (2009) identify four mechanisms that collectively interact to markedly shape CBF regulation: cerebral metabolism, ventilation, systemic cardiovascular factors (e.g., cardiac output), and sympathetic nerve activity. Importantly, this interaction is significantly impacted by exercise intensity (Ogoh & Ainslie, 2009).

Generally, CBF follows an inverted U-shaped curve during exercise (Furlong et al., 2020). With increasing exercise intensities, CBF tends to increase up to approximately 65% of one’s maximal aerobic capacity (VO2max; Furlong et al., 2020). After reaching VO2max, CBF decreases again to return to baseline levels (Furlong et al., 2020). Smith and Ainslie (2017) argue that the CBF-exercise intensity relationship may be biphasic, with CBF increasing about 10%–20% during low to moderate exercise intensities (∼60%– 70% VO2max) while plateauing or returning to baseline levels at high exercise intensities. This pattern may protect the brain from hypoperfusion and aid CA.

### 1.4 Measuring Cerebral Blood Flow

Although measuring CBF during and immediately after exercise remains challenging, several techniques have been used, with reasonable success. One of the most popular is near-infrared spectroscopy (NIRS). NIRS allows for a non-invasive, non-ionizing, low-cost, and practical evaluation of cerebrovascular responses and tissue oxygenation to exercise (Andrianopoulos et al., 2018; Auger et al., 2016). NIRS relies on near-infrared light (700- 1000 nm) to permeate skin, bone, and muscle (Simonson & Piantidosi, 1996). Continuous-wave NIRS measures changes in relative oxy- and deoxy-hemoglobin concentrations resulting from neural activity (Auger et al., 2016). Although NIRS signal is not an absolute measure of regional blood flow and can be influenced by skin vasculature signals (Auger et al., 2016; Matsukawa et al., 2015), it has the advantage of high temporal resolution while being portable and non-invasive (Shibuya et al., 2004).

Transcranial doppler ultrasonography (TCD) is an alternative that allows for the measurement of middle cerebral artery mean velocity (MCAvmean), a surrogate measure of CBF (Flück et al., 2014). TCD ultrasonography is based on the principle of the Doppler effect, whereby Doppler probe ultrasound waves penetrate the skull where moving red blood cells in the intracerebral vessels reflect the ultrasound waves (Purkayastha &Sorond, 2013). The difference in frequency between reflected and emitted waves, i.e., the Doppler shift frequency, reflects blood flow velocity (Purkayastha &Sorond, 2013). TCD measures blood flow velocity, rather than blood flow itself, as it does not consider vessel diameter (Foster et al., 2019).

Positron emission tomography (PET) is another common alternative, as it allows for a quantitative measure of global and regional CBF (Auger et al., 2016) due to its relatively high spatial resolution (Hiura et al., 2013). PET allows for measuring brain metabolism by exploring the distribution of previously injected contrast radioactive agents (Auger et al., 2016). However, the use of radioactive contrast agents makes PET a less popular option for investigating CBF in research compared to other CBF measurement techniques.

Functional magnetic resonance imaging (fMRI), unlike PET, offers fairly high temporal resolution (Logothetis, 2008), and can detect changes in cerebral blood oxygenation and flow that co-occur with neural activity. It is has become a very common measurement tool in cognitive neuroscience research (Logothetis, 2008). Like all hemodynamic -based modalities, fMRI measures a surrogate signal that is susceptible to physical and biological constraints, such as neural mass activity (Logothetis, 2008). Compared to fNIRS, fMRI lacks portability and is less practical for longitudinal monitoring (Chen et al., 2020).

Finally, arterial-spin labelling (ASL) magnetic resonance imaging is a non-invasive, quantitative method that relies on magnetically labeled arterial blood water protons as endogenous tracers to measure tissue perfusion (Steventon et al., 2020). ASL directly measures CBF by labeling inflowing water to tissue (Foster et al., 2019).

### 1.5. Present Review

The literature on the effect of acute cardiovascular exercise on CBF has not received sufficient attention, especially given that the determinants and moderators affecting differences within and across individuals remain poorly understood. To address this gap, we conducted a systematic review of the acute effect of cardiovascular exercise on CBF, measured via diverse neuroimaging techniques and across a range of exercise characteristics.

## 2. Methods

### 2.1. Protocol and Registration

We adhered to the Preferred Reporting Items for Systematic Reviews and Meta-Analyses (PRISMA) checklist (Moher et al., 2015) to conduct this review. Before beginning the review, we registered our protocol in accordance with PRISMA with the international prospective register of systematic reviews (PROSPERO; CRD42022299841). Registration of the protocol was completed on February 24, 2022.

### 2.2. Information Source and Search strategy

The systematic literature search was performed on the following seven electronic databases: PsychINFO (all fields), Web of Science (all fields), Science Direct (keywords, title, author), ProQuest (all fields), PubMed (all fields), SPORTDiscus (all fields), and Scopus (title, abstract, keywords) in February 2022. Additionally, Google Scholar, PsyArXiv, and bioRxiv were searched to identify potential gray literature. Search terms are shown in Table 1.

**Table 1.**
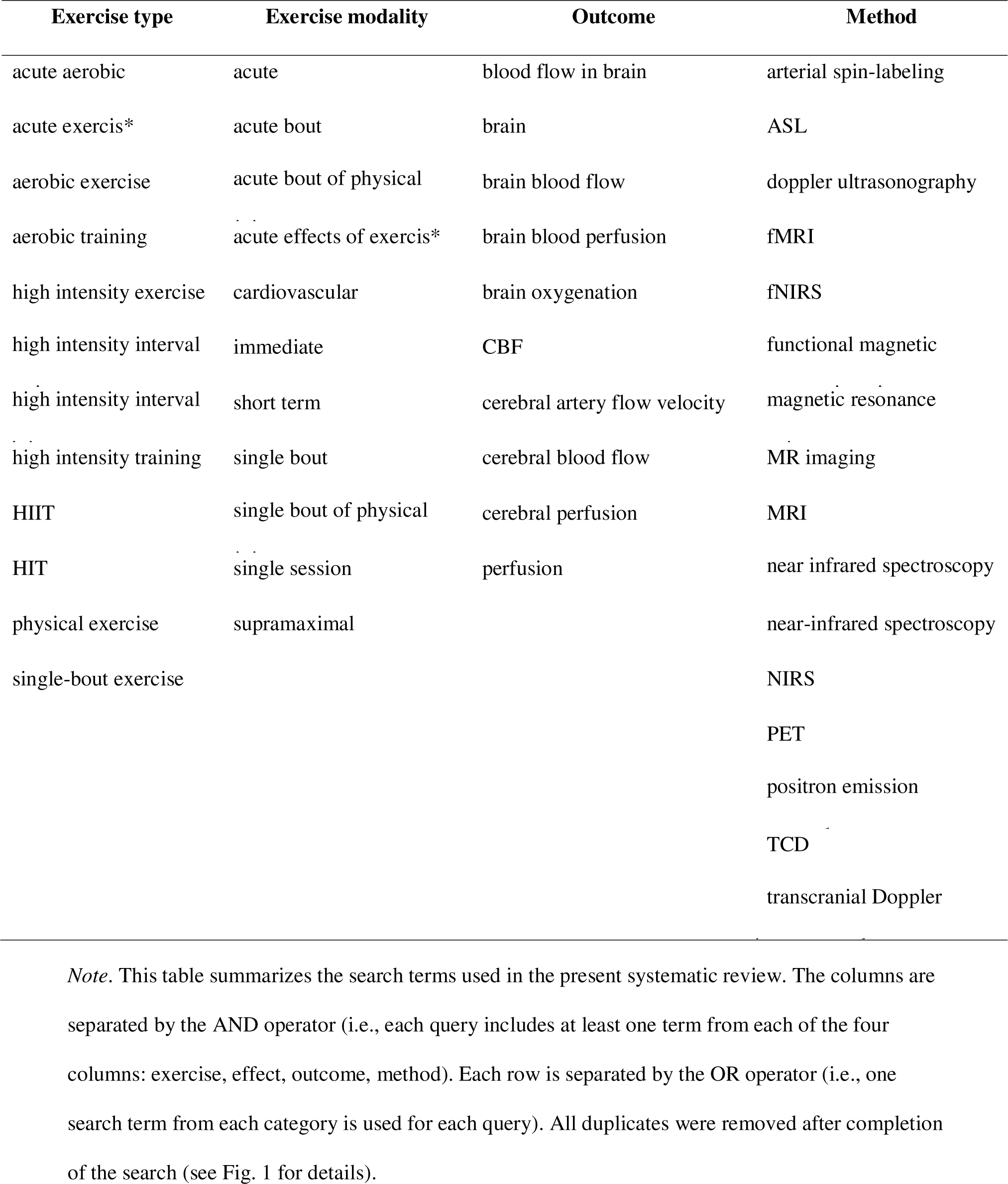
Search Terms

| **Exercise type** | **Exercise modality** | **Outcome** | **Method** |
| --- | --- | --- | --- |
| acute aerobic | acute | blood flow in brain | arterial spin-labeling |
| acute exercis* | acute bout | brain | ASL |
| aerobic exercise | acute bout of physical | brain blood flow | doppler ultrasonography |
| aerobic training | acute effects of exercis* | brain blood perfusion | fMRI |
| high intensity exercise | cardiovascular | brain oxygenation | fNIRS |
| high intensity interval | immediate | CBF | functional magnetic |
| high intensity interval | short term | cerebral artery flow velocity | magnetic resonance |
| high intensity training | single bout | cerebral blood flow | MR imaging |
| HIIT | single bout of physical | cerebral perfusion | MRI |
| HIT | single session | perfusion | near infrared spectroscopy |
| physical exercise | supramaximal |  | near-infrared spectroscopy |
| single-bout exercise |  |  | NIRS |
|  |  |  | PET |
|  |  |  | positron emission |
|  |  |  | TCD |
|  |  |  | transcranial Doppler |

*Note.* This table summarizes the search terms used in the present systematic review. The columns are separated by the AND operator (i.e., each query includes at least one term from each of the four columns: exercise, effect, outcome, method). Each row is separated by the OR operator (i.e., one search term from each category is used for each query). All duplicates were removed after completion of the search (see Fig. 1 for details).

### 2.3. Eligibility Criteria

We used PICOS to screen relevant studies (Richardson et al., 1995). “PICOS” is short for participants (P), intervention (I), comparisons (C), outcomes (O), and study design (S) (Richardson et al., 1995). The present systematic review focused on studies featuring human participants only (P). The included samples encompass human participants regardless of their age, fitness level, or medical history. Studies focusing on acute aerobic or high-intensity exercise as the independent variable (I) were included. No specific restrictions for comparisons (C) were applied. The outcomes (O) of interest include potential changes in cerebral blood flow measured with arterial spin-labeling (ASL), functional magnetic resonance imaging (fMRI,), positron emission tomography (PET), transcranial Doppler ultrasonography (TCD), or near-infrared spectroscopy (NIRS) assessments (results of such measures were required to be used as the dependent variable). Study design (S) criteria included publication in a peer-reviewed journal, in English, and including one of the following designs: experimental, correlational, empirical, group, and within-subject, with effect sizes or the necessary information to calculate effect sizes.

The search yielded 483 results. After removing duplicates and retrieving articles, 224 studies were full-text screened for inclusion and exclusion criteria (see Fig. 1 for a detailed flow chart). A total of 172 studies were excluded as they did not meet our inclusion criteria. The most common reasons for exclusion were: a focus on cognitive functioning rather than cerebral perfusion, exercise interventions that were chronic rather than acute, a focus on hypoxia, and missing information (e.g., effect sizes). Fifty-two articles met all eligibility criteria and were therefore included in the present review.

**Figure 1.**
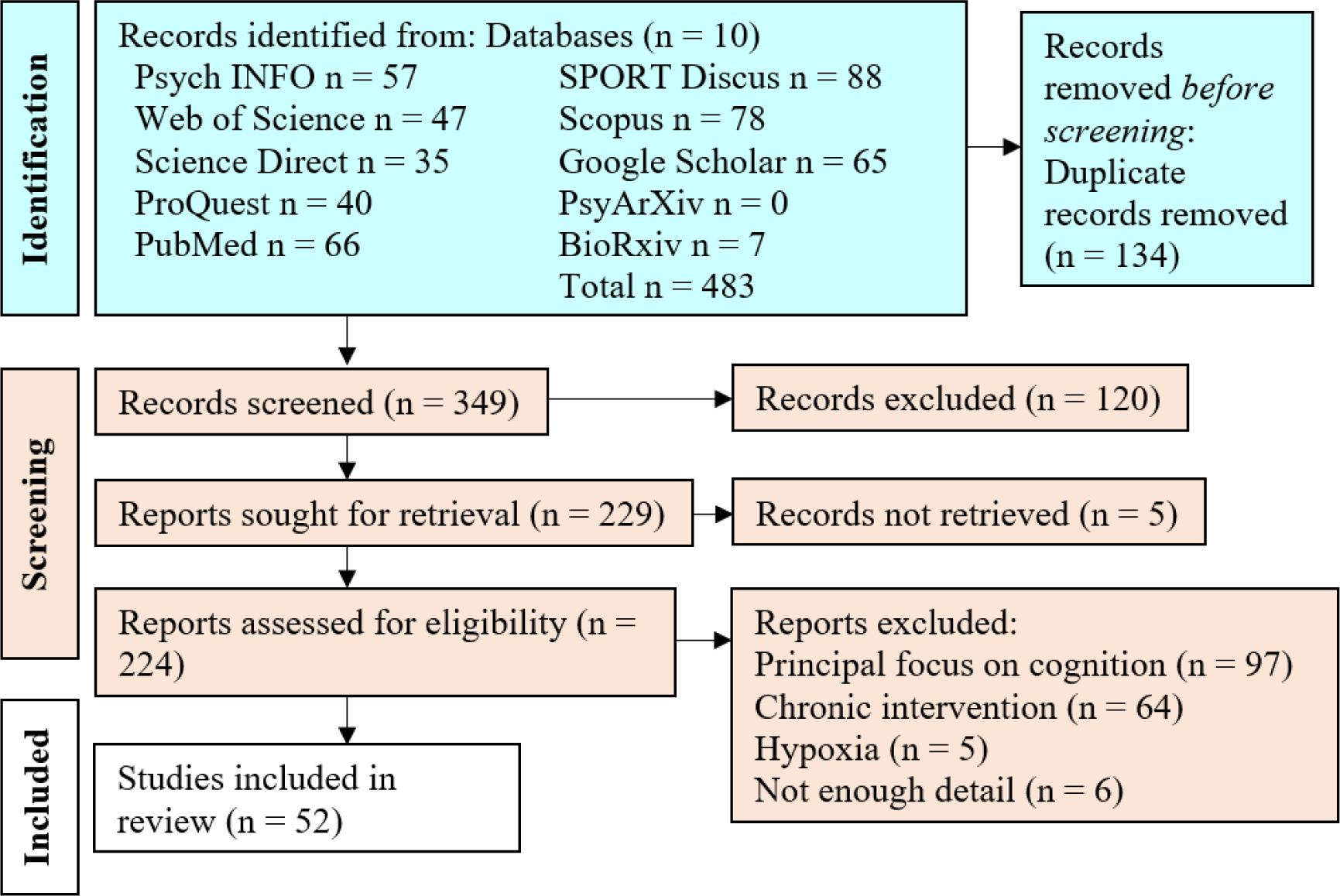
PRISMA flow chart

### 2.4. Data Extraction

We extracted information about the authors, year of publication, population characteristics, intervention characteristics (e.g., aerobic or high-intensity exercise), measures used to investigate cerebral perfusion (e.g., ASL, PET, fNIRS, (f)MRI, or TCD), and effect sizes or information to calculate effect sizes (e.g., group means and standard deviations). Moreover, we extracted information about the main findings for each included study.

### 2.5. Risk of Bias Assessment

We assessed the risk of bias in each of the 52 studies using the Cochrane Collaboration’s Risk of Bias tool (RoB 2; Sterne et al., 2019). Each study was rated in five categories, including the “randomization process”, “deviations from the intended interventions,” “selection of the reported result,” “missing outcome data “, and “measurement of the outcome” (Sterne et al., 2019). An overall risk of bias score was determined based on each paper’s individual scores in each of the five categories. The risk of bias was rated as either “high,” “some concerns,” or “low” (Sterne et al., 2019).

## 3. Results

Participant and protocol summary statistics for the studies included in this systematic review are presented Table 2 and Table 3, respectively. We present results in more details hereafter, including via assessments of overall and specific bias, as well as with study-level results.

**Table 2.** Participants and Study Characteristics

| **Participant characteristics** |  | **N** | **%** |
| --- | --- | --- | --- |
| **Age** | ≤ 15 yrs. | 81 | 6.89 % |
|  | 16-17 yrs. | 51 | 4.34 % |
|  | 18-29 yrs. | 422 | 35.95 % |
|  | 30-49 yrs. | 190 | 16.18 % |
|  | ≥ 50 yrs. | 358 | 30.50 % |
|  | Other | 72 | 6.14 % |
|  | **Total** | **1174** | **100 %** |
| **Sex** | male | 727 | 61.93 % |
|  | female | 379 | 32.29 % |
|  | not reported | 68 | 5.79 % |
|  | **Total** | **1174** | **100 %** |
| **Health Status** | no relevant condition | 899 | 76.58 % |
|  | presence of relevant condition | 275 | 23.42 % |
|  | **Total** | **1174** | **100 %** |
| **Presence of** | IDC | 77 | 28 % |
|  | COPD | 52 | 18.91 % |
|  | VHD | 33 | 12 % |
|  | BD | 31 | 11.27 % |
|  | Stroke | 36 | 13.09 % |
|  | HD gene | 19 | 6.91 % |
|  | HF | 18 | 6.55 % |
|  | CFS | 6 | 2.18 % |
|  | high CVR | 3 | 1.09 % |
|  | **Total** | **275** | **100 %** |
| **Fitness Level** | trained | 85 | 7.24 % |
|  | active | 221 | 18.82 % |
|  | sedentary/ not assessed | 868 | 73.94 % |
|  | **Total** | **1174** | **100 %** |

**Table 3.**
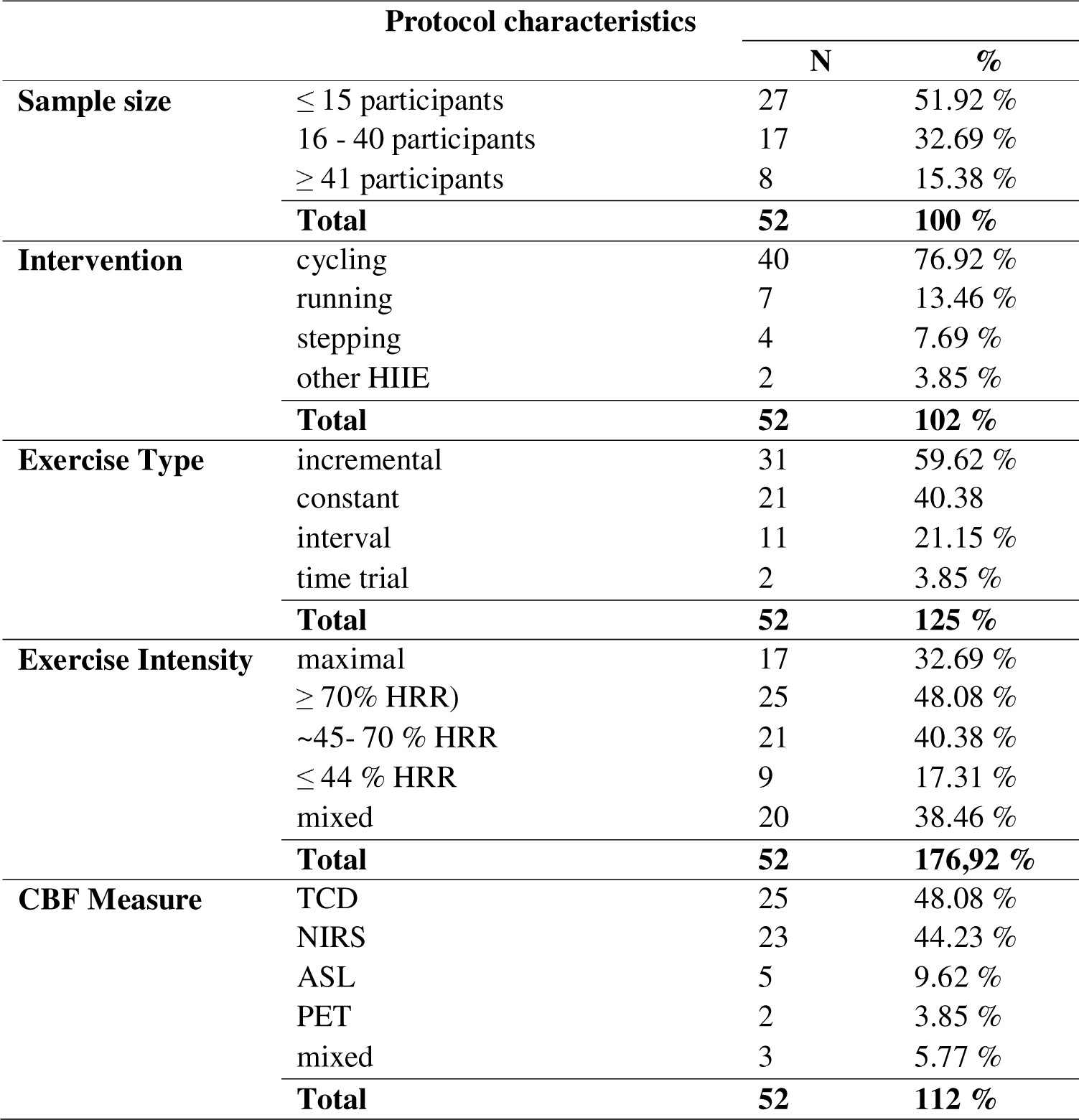

### 3.1. Risk of Bias

Table 4 shows the results of the Risk of Bias assessment. No study was rated as having a “high” overall risk of bias. Ten studies were deemed to have “some concerns” regarding risk of bias, whereas 42 studies received an overall risk of bias rating of “low.” Randomization was rated as “some concern” more frequently than other domains, either due to studies not applying randomization and/or blinding practices, or to procedures not being described in sufficient detail to assess randomization.

**Table 4.**
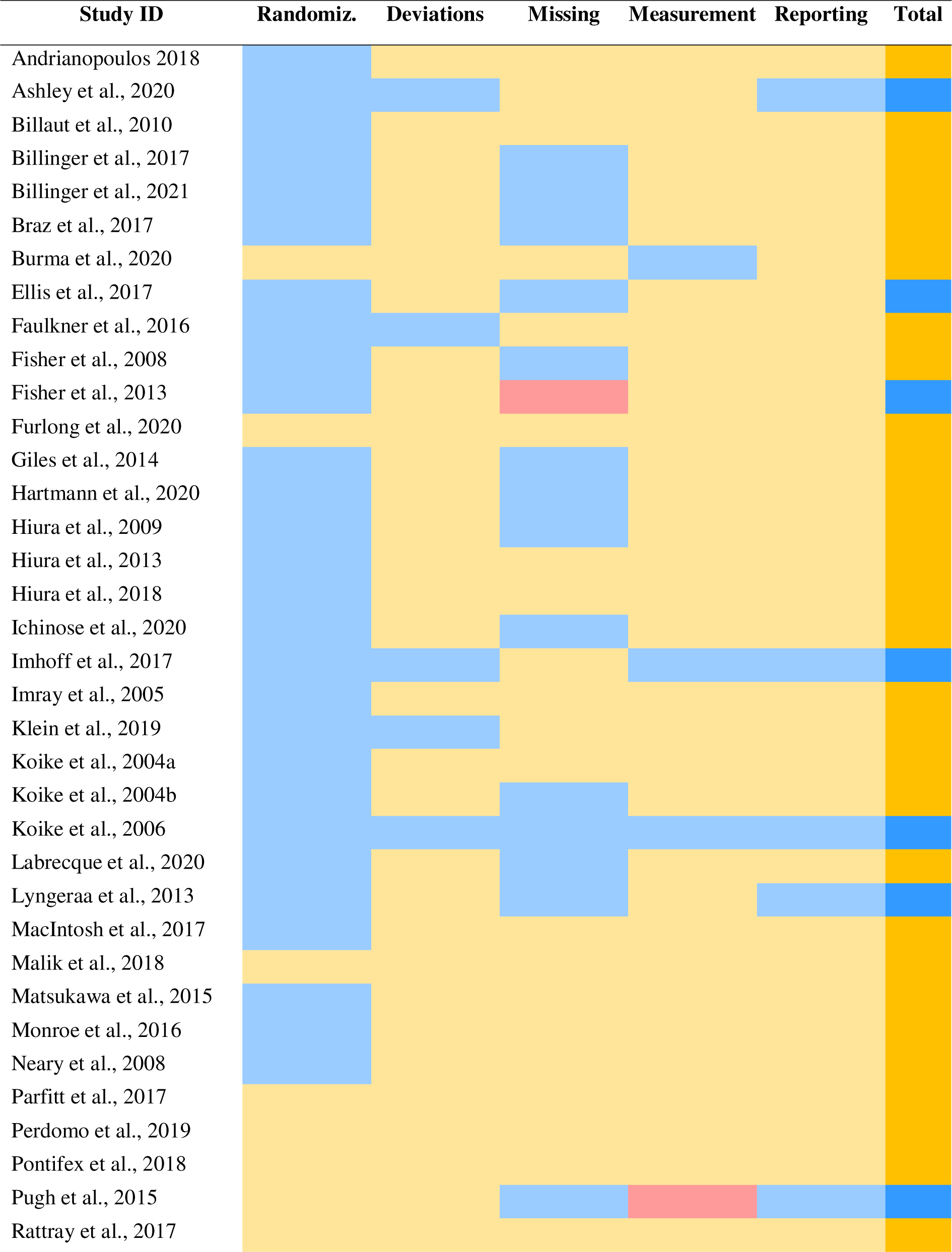
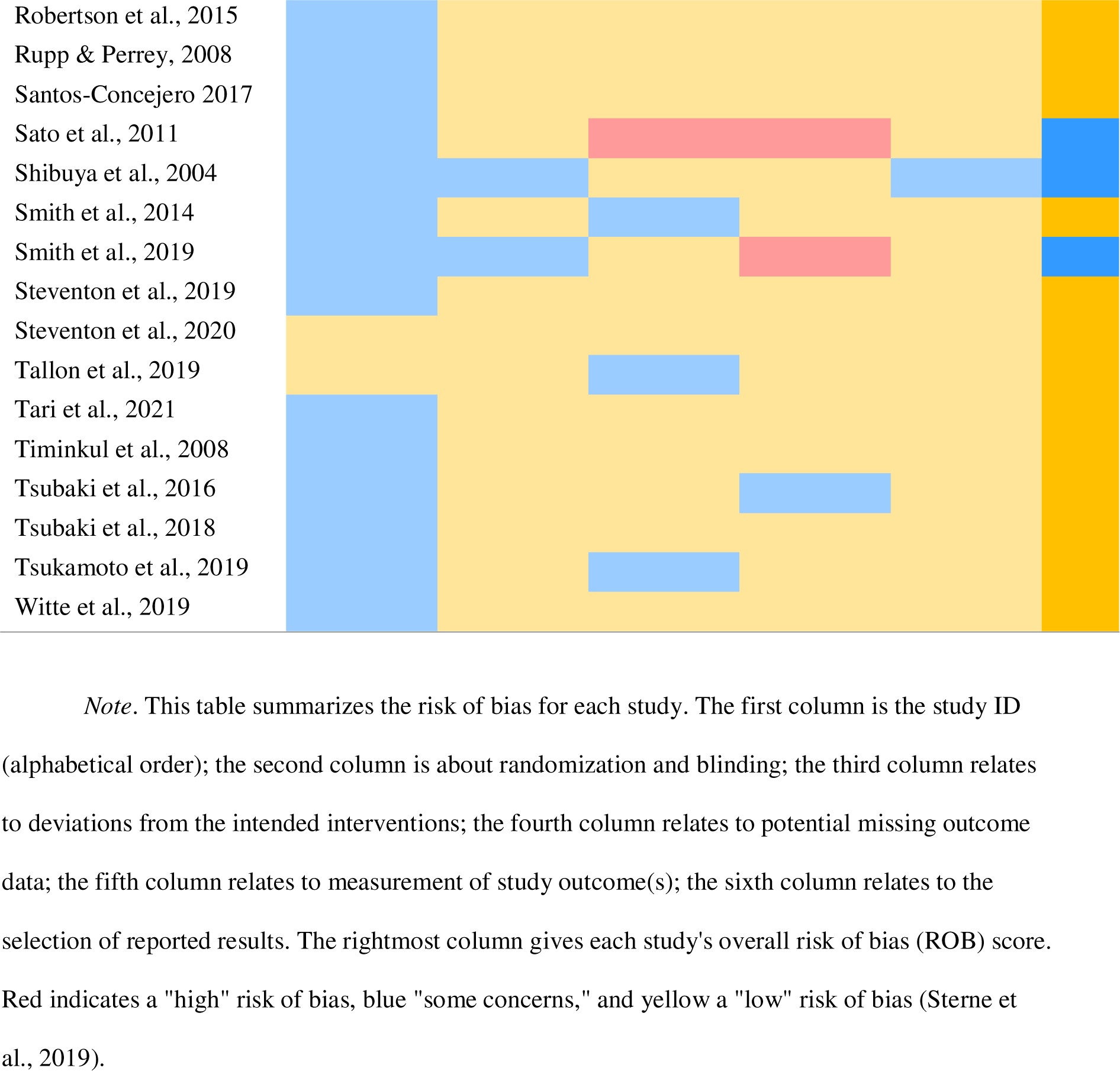
Risk of Bias (ROB) Assessment Across All Included Studies

### 3.2. Participant Characteristics

This systematic review includes peer-reviewed studies that investigated the effects of acute cardiovascular exercise on cerebral blood flow in children (0 - 15 years, n = 81), adolescents (16-17 years, n = 51), young adults (18-29 years, n = 422), adults (30-49 years, n = 190), older adults (≥ 50 years, n = 358), and others (n =72). The most common age group represented in this systematic review are young adults aged between 18 and 29 years. Next, 30.50 % of the total sample are adults aged 50 years and over, whereas adults aged between 30 and 49 years represent 16.18 % of the overall sample. Four studies (Ellis et al., 2017; Malik et al., 2018; Pontifex et al., 2018; Tallon et al., 2019) included participants aged 15 years or younger, whereas a single study focused on adolescents aged between 16 and 17 years (MacIntosh et al., 2017). Note that two studies were considered unclear in reporting age-related information. For example, Hartmann and colleagues (2020) state that their sample includes 54 smokers and non-smokers divided into a “young” (p.2) (22 (±1.6) years) and a “middle-aged” (p.2) (33 years (±7.8) group; however it remains unclear how many participants were included in each category. Similarly, Ichinose and colleagues (2020) did not mention the age of their 18 participants. This is unfortunate as age may significantly impact the exercise-CBF relationship. Some studies reported significant CBF by age correlations (Braz et al., 2017; Ellis et al., 2017; Fisher et al., 2013; Klein et al., 2019), whereby older participants tend to show lower cerebral perfusion compared to their younger counterparts. However, Ellis and colleagues (2017) showed that the increase in CBF in the anterior cerebral circulation during exercise in children was approximately half of that observed in adults. In contrast, Fisher and colleagues (2008) reported no CBF by age effect.

Apart from age, sex could also impact the effect cardiovascular exercise has on CBF. Of the 1,174 participants collectively included in this systematic review, 727 (61.9 %) identified as male, 379 as female (32.29 %), whereas the sex of 68 (5.79 %) was not reported. Sex may impact the CBF response to exercise; for example, Ashley and colleagues (2020) found that the cerebral vascular conductance index (CVCi) was significantly greater in females compared to males in their sample, while the cerebral perfusion pressure (CPP) was significantly greater in males compared to females.

Health and fitness also play a significant role in the relationship between exercise and CBF. To investigate the effect of these factors, we grouped participants based on the existence or absence of conditions known to impact the effect cardiovascular exercise may have on CBF. Therefore, the label ‘healthy’ does not describe the absence of any medical condition but rather indicates that participants had no known illness, disability, or impairment known to affect a potential exercise-induced effect on CBF. The majority of the participants included in this review met the ‘healthy’ criteria, while the remaining 23.4 % of participants disclosed a condition that may be relevant to the effect of cardiovascular exercise on cerebral perfusion. Of those, the most common condition was Idiopathic Dilated Cardiomyopathy (IDC, n = 77). Furthermore, 52 presented with clinically stable Chronic Obstructive Pulmonary Disease (COPD), 33 had valvular heart disease (VHD), 31 presented with bipolar disorder (BD), 36 survived a stroke, 19 participants were Huntington’s Disease gene-positive, 18 participants experienced heart failure with 9 of them carrying a left ventricular assist device (LVAD), 6 participants experienced chronic fatigue syndrome (CFS), and 3 participants were deemed at high cardiovascular risk. MacIntosh and colleagues (2017) noticed that participants with BD were characterized by an increased CBF in the left medial frontal gyrus and bilateral middle cingulate regions at baseline compared to participants without BD. Furthermore, participants with BD exhibited a more considerable CBF decrease fifteen minutes post-exercise (MacIntosh et al., 2017). Similarly, Neary and colleagues (2008) reported reduced cerebral oxygenation in participants with CFS compared to controls. Hartmann and colleagues (2020) found that non-smokers exhibited a larger increase in gCBF during exercise compared to smokers. Finally, Steventon and colleagues (2020) reported that participants who carry a positive gene for Huntington’s Disease showed a significant increase in rCBF, while controls did not. Besides health, fitness may play an essential role in the effects of acute exercise on CBF (Braz et al., 2017).

With respect to fitness level, participants were characterized as either trained (n = 85), active (n = 221), or sedentary/ not assessed (n = 868). The difference between trained and active participants was that trained participants were athletes that regularly competed in their sport, while active participants exercised regularly in a recreational setting. However, the most common group making up 73.9 % of the total sample, were sedentary participants or participants whose fitness level had not been reported. Unreported fitness levels could potentially cloud results as fitness seems to play a crucial role in the effect of cardiovascular exercise on CBF (Ellis et al., 2017). For example, Braz and colleagues (2017) found that the bilateral ICA blood flow was 31% higher in trained compared to untrained individuals. Smith and colleagues (2014) highlight that the altitude at which an exercise is performed significantly shapes the effect that cardiovascular exercise can have on CBF including recovery. In particular, Smith and colleagues (2014) report increased gCBF during exercise and recovery at high altitude (HA) compared with sea level (SL).

### 3.3. Exercise Characteristics

First, included studies focused on various exercise interventions including cycling (electronically braked cycle ergometer; n = 40), running (treadmill; n = 7), stepping exercise (recumbent stepper; n = 4), and other HIIE (n = 2). Note that one study used both treadmill-running and cycling on an electronically braked cycle ergometer as exercise intervention to test the effects of exercise on cerebral perfusion (Furlong et al., 2020). For example, Sato and colleagues (2011) observed a significant increase in MCA V_mean_ from rest to 80% VO2peak followed by a significant decrease in MCA V_mean_ from 80% VO2peak during a cycling intervention. Second, the exercise interventions consisted predominantly of incremental exercise (n = 31), followed by constant (n = 21), interval (n = 11), and time trial interventions (n = 2). Furthermore, thirteen studies employed mixed exercise interventions (e.g., one constant and one incremental running trial). Third, exercise intensities were reported as maximal (n = 17), high (≥ 70% HRR; Andrianopoulos et al., 2018) (n = 25), moderate (∼45- 70 % HRR; Hiura et al., 2018; Robertson et al., 2015) (n = 21), or low (≤ 44 % HRR; Hiura et al., 2018; Robertson et al., 2015) (n = 9). Furthermore, twenty studies included several exercise intensities. Tsukamoto and colleagues (2019) concluded that cerebral autoregulation (CA) during HIIE remains stable. On the other hand, Burma and colleagues (2020) argue that CA is impaired following both moderate continuous exercise and HIIT. Others (Billinger et al., 2021; Robertson et al., 2015) state that low-intensity exercise does not affect the cerebrovascular response (CVR).

### 3.4. Cerebral Blood Flow Measurement Characteristics

As summarized in Table 3, the most common measure of cerebral perfusion used in the included studies was Transcranial Doppler Ultrasound (TCD; n =25), followed by Near-infrared spectroscopy (NIRS; n = 23), Arterial spin labeling (ASL; n = 5), and Positron Emission Tomography (PET; n = 2). Three studies used TCD and NIRS to determine cerebral perfusion (Imhoff et al., 2017; Imray et al., 2005; Tari et al., 2021).

TCD data suggests that cerebral blood flow may significantly increase as a result of acute aerobic exercise (Billinger et al., 2017; Burma et al., 2020; Ellis et al., 2017; Fisher et al., 2008; Fisher et al., 2013; Furlong et al., 2020; Imhoff et al., 2017; Klein et al., 2019; Labrecque et al., 2020; Lyngeraa et al., 2013; Parfitt et al., 2017; Perdomo et al., 2019; Pugh et al., 2015; Rattray et al., 2017; Sato et al., 2011; Smith et al., 2019; Witte et al., 2019), with only a single study showing that CBF remained unchanged (Burma et al., 2020). Specifically, 17 papers reported a significant increase in middle cerebral artery blood flow mean velocity (MCAV) (Billinger et al., 2017; Burma et al., 2020; Ellis et al., 2017; Fisher et al., 2008; Fisher et al., 2013; Furlong et al., 2020; Imhoff et al., 2017; Klein et al., 2019; Labrecque et al., 2020; Lyngeraa et al., 2013; Parfitt et al., 2017; Perdomo et al., 2019; Pugh et al., 2015; Rattray et al., 2017; Sato et al., 2011; Smith et al., 2019; Witte et al., 2019). Additionally, Labrecque and colleagues (2020) and Pugh and colleagues (2015) reported a significant increase in posterior cerebral artery mean blood velocity (PCAv_mean_) during exercise. Sato and colleagues (2011) highlighted a significant increase in common carotid artery (CCA) and vertebral artery (VA) blood flow from rest to 80% VO2 peak during a cycling intervention, whereas Burma and colleagues (2020) reported that CBF velocity in the middle cerebral artery (MCA) and the posterior cerebral artery (PCA) remained unchanged during moderate-intensity continuous cycling and high-intensity interval cycling.

NIRS data underlines the complex nature of changes in CBF associated with acute aerobic exercise. First, ten studies reported a significant increase in the concentration of oxyhemoglobin (O2Hb) in the brain during acute aerobic exercise (Giles et al., 2014; Ichinose et al., 2020; Koike et al., 2004a; Koike et al., 2004b; Koike et al., 2006; Rupp & Perrey, 2008; Malik et al., 2018; Monroe et al., 2016; Timinkul et al., 2008; Tsubaki et al., 2018). Interestingly, Koike and colleagues (2004a; 2004b; 2006) highlight that a significant increase in the O2Hb concentration could only be detected in the healthy control groups. Second, participants with IDC (Koike et al., 2004a; Koike et al., 2006) or VHD (Koike et al., 2004b) showed a significant decrease in O2Hb during exercise. Third, the concentration of deoxyhemoglobin (dHb) significantly increased in healthy controls and participants with VHD (Ichinose et al., 2020; Koike et al., 2004b; Malik et al., 2018; Monroe et al., 2016; Rupp & Perrey, 2008; Timinkul et al., 2008), whereas it remained unchanged in some healthy controls (Koike et al., 2004b), and significantly decreased in recreationally active students towards the end of cycling exercises of varying intensities (Tari et al., 2021). Fourth, three studies explicitly reported an increase in the total blood volume (tHb) in the cerebral tissue in patients with COPD (Andrianopoulos et al., 2018) and participants with no known relevant conditions (Giles et al., 2014; Rupp & Perrey, 2008). Fifth, Timinkul and colleagues (2008) observed a significant increase in the tissue oxygenation index (TOI) in some participants, whereas TOI remained unchanged in others. On the other hand, Neary and colleagues (2008) and Santos-Concejero and colleagues (2017) report a significant decrease in TOI. Lastly, Faulkner and colleagues (2016) note a significant increase in mean regional cerebral tissue oxygen saturation (mean rSO2).

Regardless of the specific CBF measurement technique used, most studies reported an increase in global CBF during exercise (Hartmann et al., 2020; Hiura et al., 2009; Hiura et al., 2018; Perdomo et al., 2019; Smith et al., 2014; Tari et al., 2021). Regional cerebral blood flow (rCBF), on the other hand, may increase during (Hiura et al., 2013; Hiura et al., 2018; Lyngeraa et al., 2013; Robertson et al., 2015; Steventon et al., 2019; Steventon et al., 2020) or decrease (Hiura et al., 2018; MacIntosh et al., 2017; Pontifex et al., 2018; Robertson et al., 2015; Robertson et al., 2015) after a single bout of exercise. Specifically, Hiura and colleagues (2018) reported a significant increase in cerebral blood flow in the following areas: midbrain, pons, medulla, cerebellar hemispheres and vermis, sensorimotor cortex for the bilateral legs (M1Leg and S1Leg), right caudal nucleus, right and left parietal operculum, insular cortex, and right anterior and posterior cingulate gyrus. Meanwhile, Steventon and colleagues (2019) found a significant increase in hippocampal CBF during exercise and up until 55–65 minutes after exercise cessation. Robertson and colleagues (2015) highlight that the increase in rCBF was confined to the moderate-intensity condition. Moreover, Steventon and colleagues (2020) argue that the reported increase in rCBF was only observed in participants that are Huntington’s Disease gene-positive. On the other hand, Hiura and colleagues (2018) noted a significant decrease in rCBF in the cerebellum, the inferior and superior temporal gyrus, the left thalamus, the pons, and the left piriform lobe ten minutes after exercise cessation. After completing a twenty-minute running exercise, participants in Pontifex and colleagues’ (2018) study exhibited a significant decrease in cerebral oxygenation in the left inferior frontal gyrus, extending across the lateral prefrontal cortex, superior temporal gyrus, planum temporale, insular cortex, angular gyrus, inferior temporal cortex, and clusters in the anterior cingulate cortex and bilateral insular cortices. MacIntosh and colleagues (2017) measured a significant decrease in cerebral perfusion confined to the left precentral gyrus, left superior frontal gyrus, left occipital pole, right precentral gyrus, left postcentral gyrus, and the left frontal pole in participants with BD approximately fifteen minutes post- exercise. Furthermore, Robertson and colleagues (2015) report a significant decrease in cerebral perfusion in the right lentiform nucleus and the left superior and middle temporal gyri thirty to fifty minutes after completing the cycling exercise. Looking at the cerebrovascular response (CVR) changes during exercise, several studies point out that cerebral perfusion may follow an inverted U response, whereby CBF increases at the onset of exercise, then plateaus, and finally decreases towards the peak or end of the exercise (Billaut et al., 2010; Imray et al., 2005; Matsukawa et al., 2015; Neary et al., 2008; Pugh et al., 2015; Santos-Concejero et al., 2017; Sato et al., 2011; Shibuya et al., 2004; Tallon et al., 2019; Tsubaki et al., 2016). Study-level results are presented in Table 5.

**Table 5.**
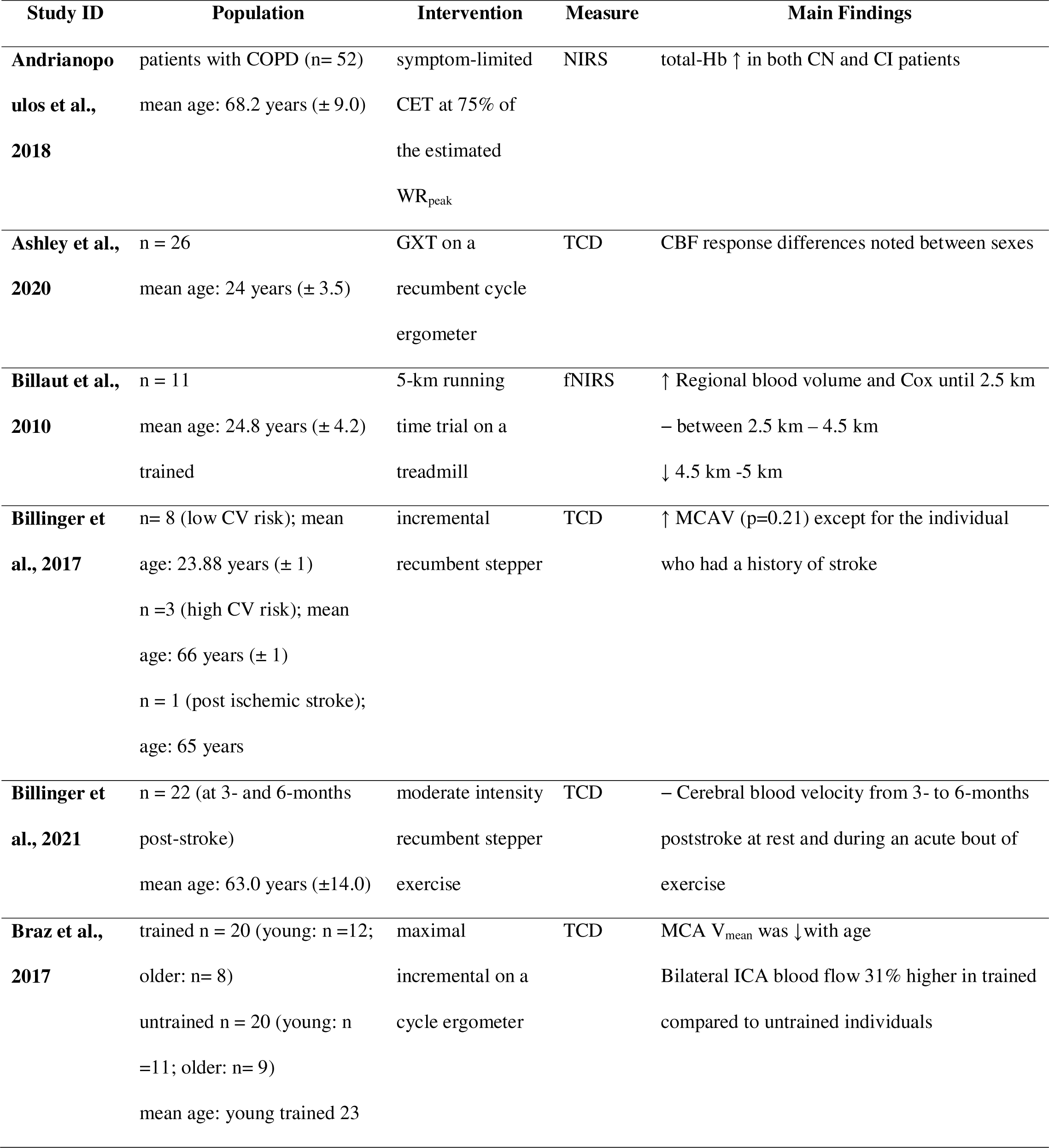
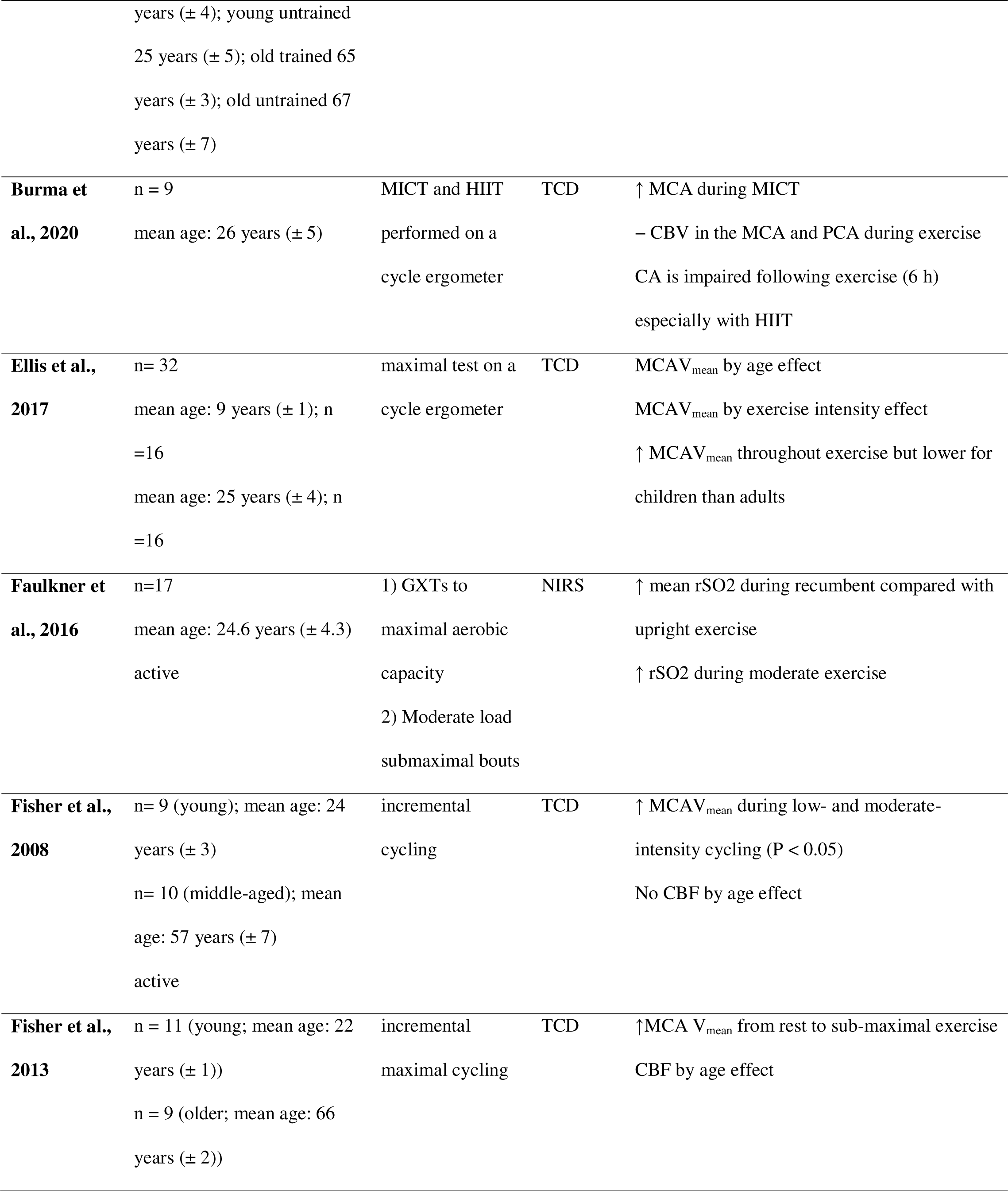
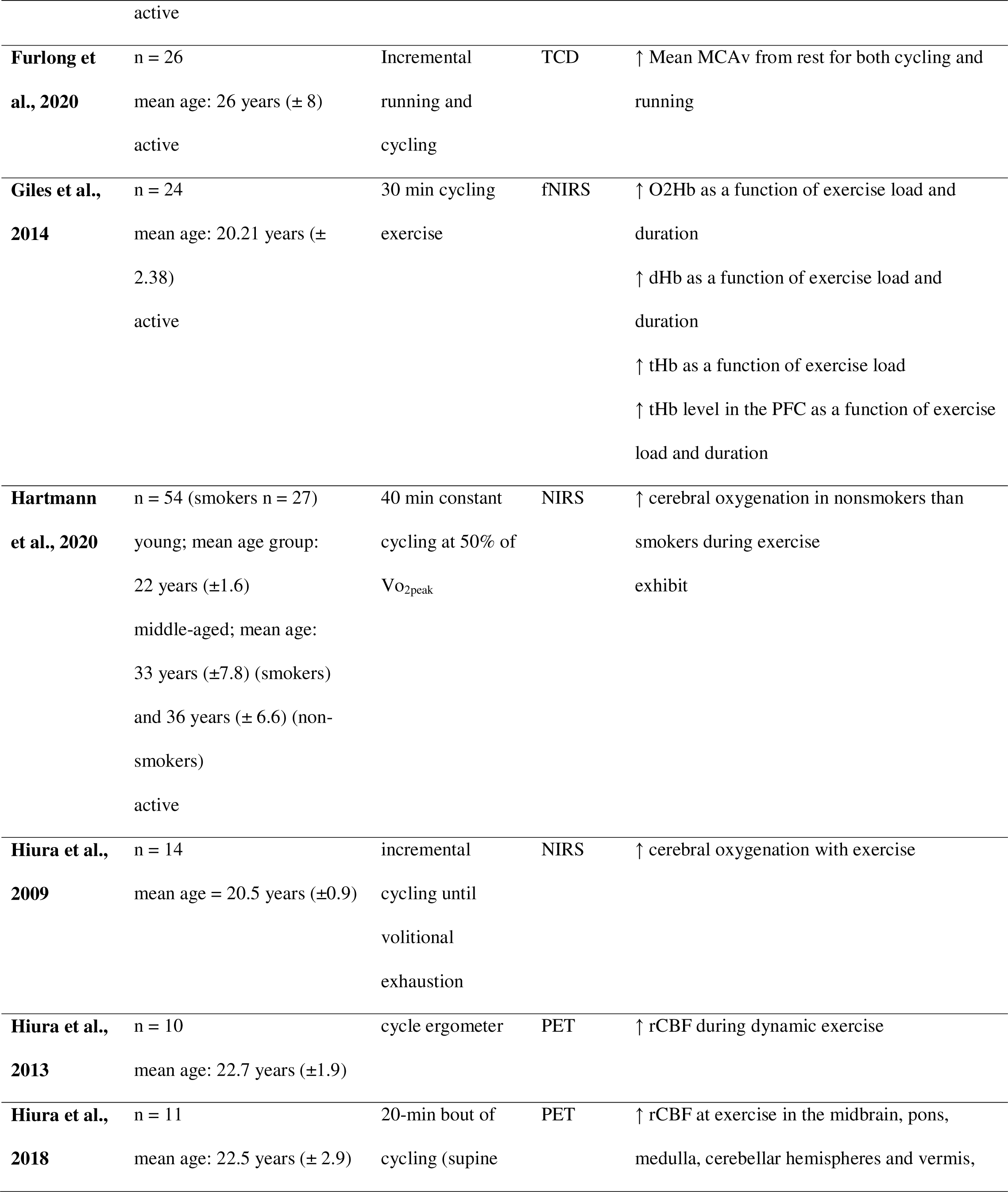
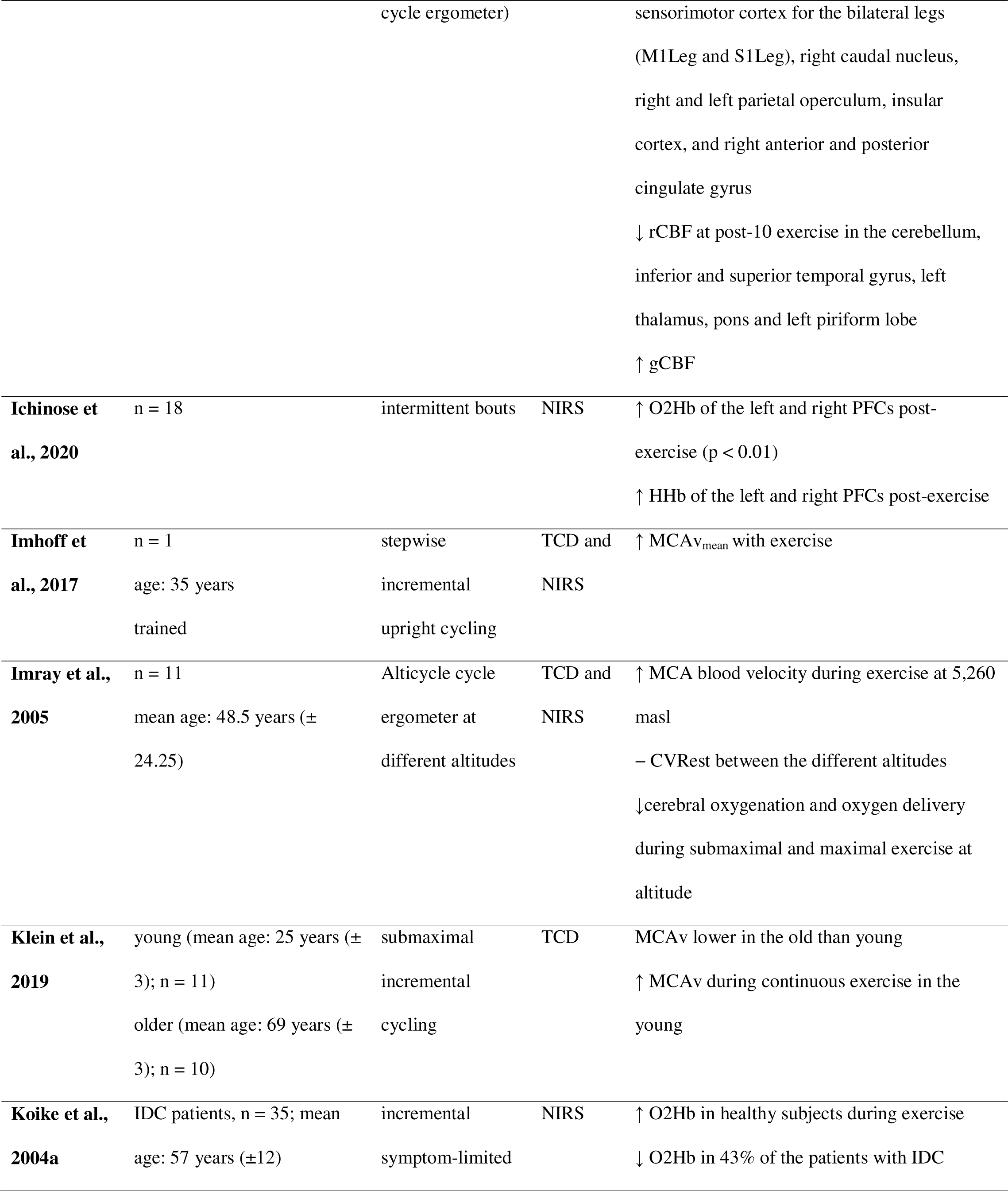
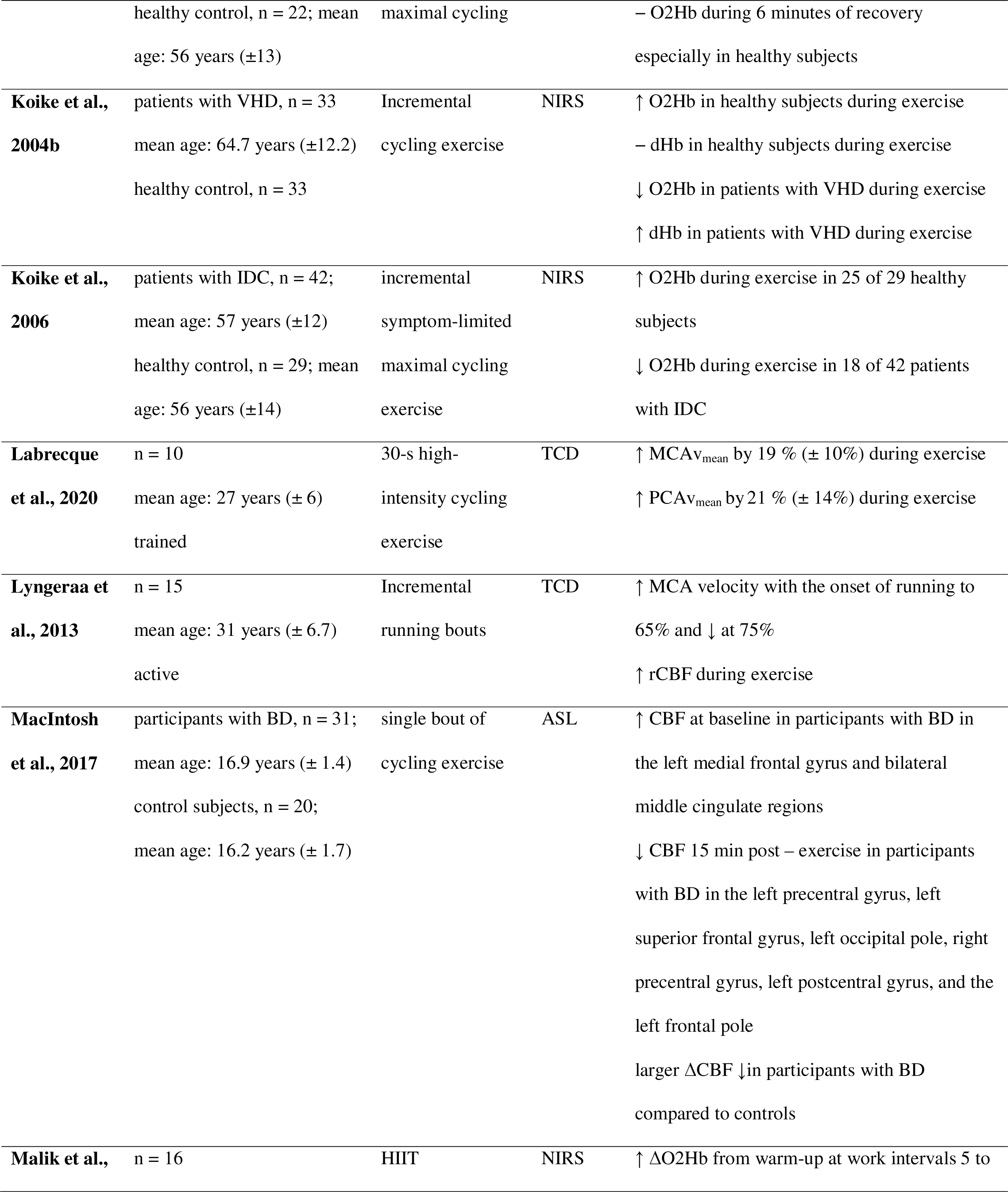
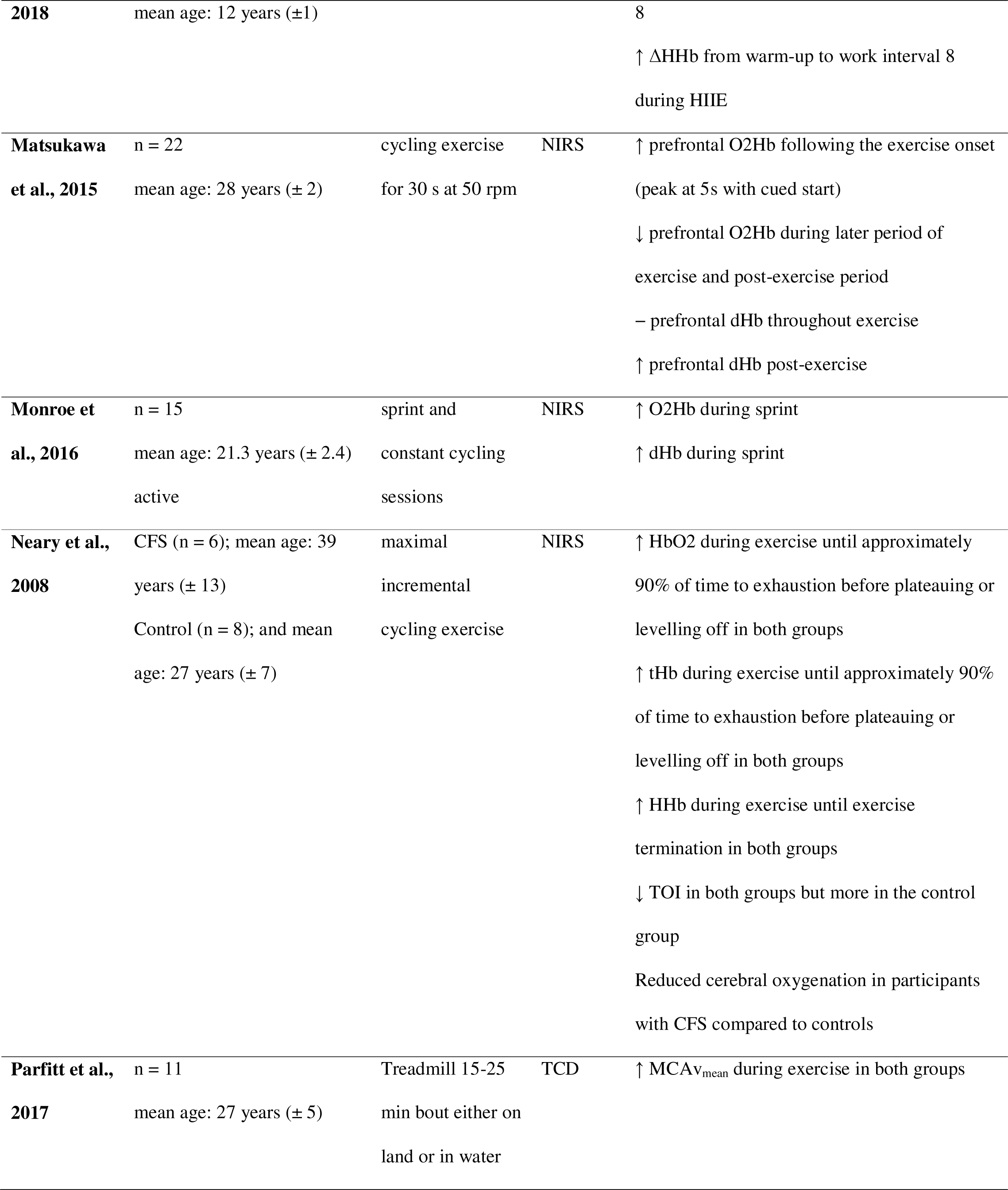
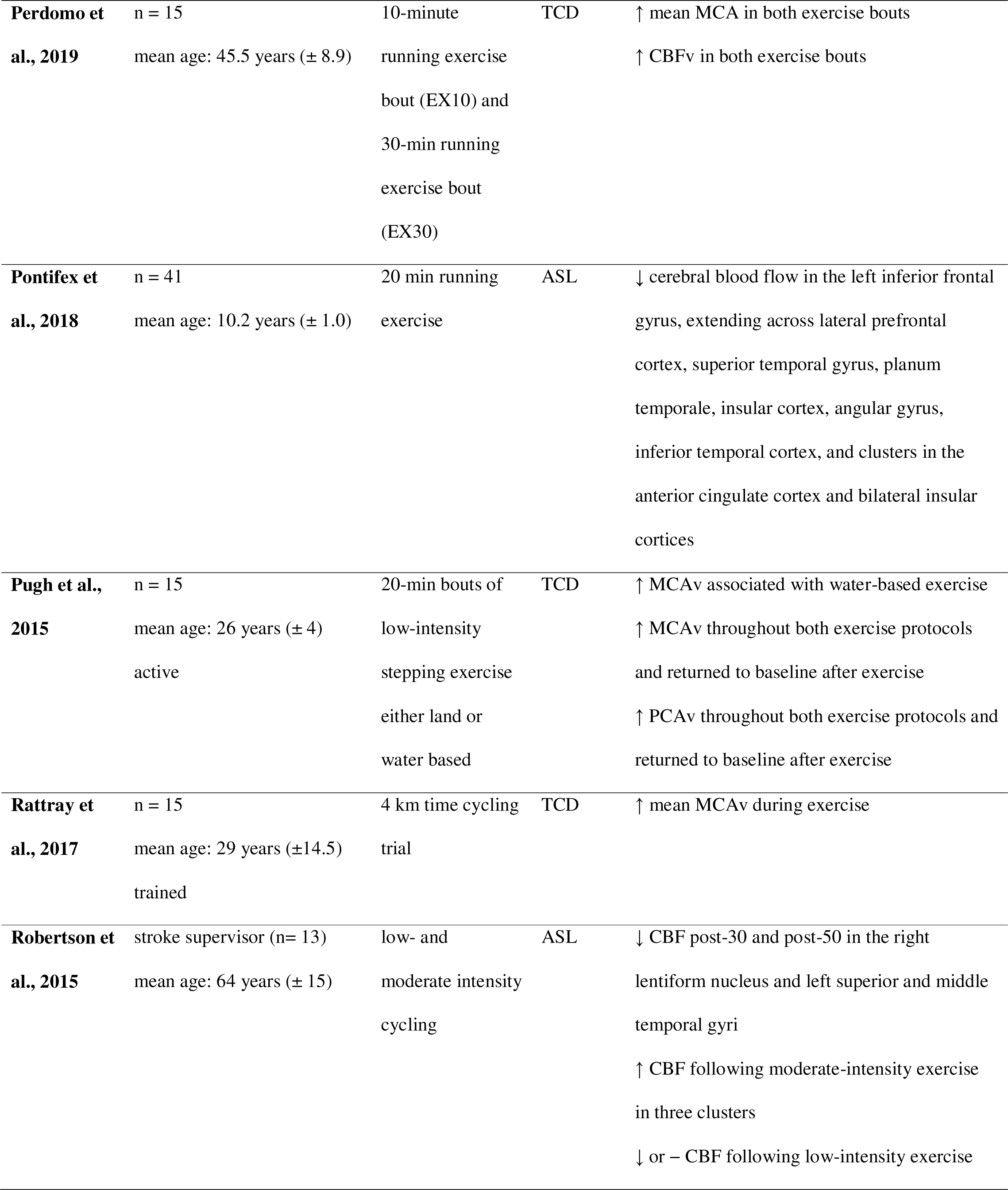
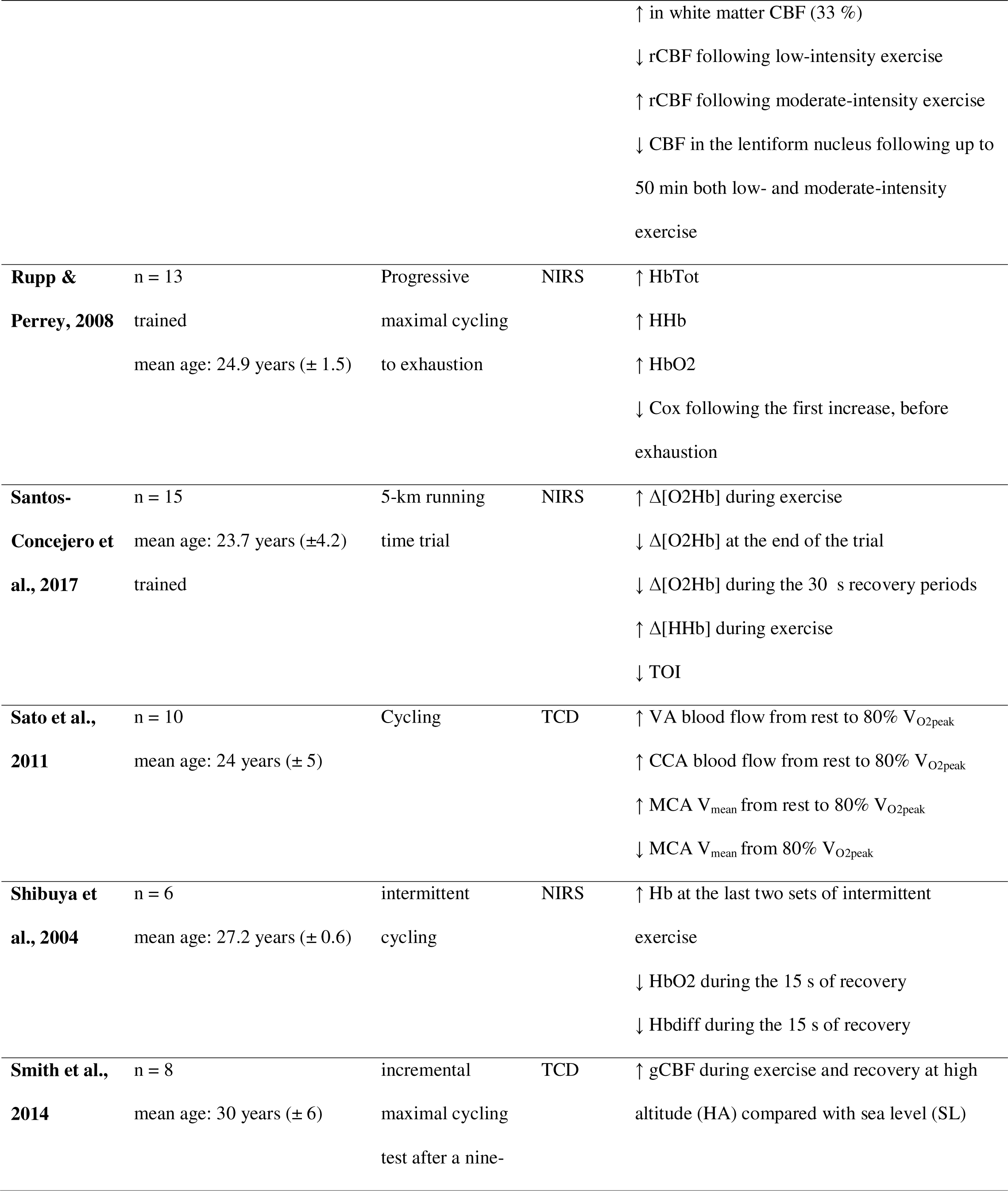
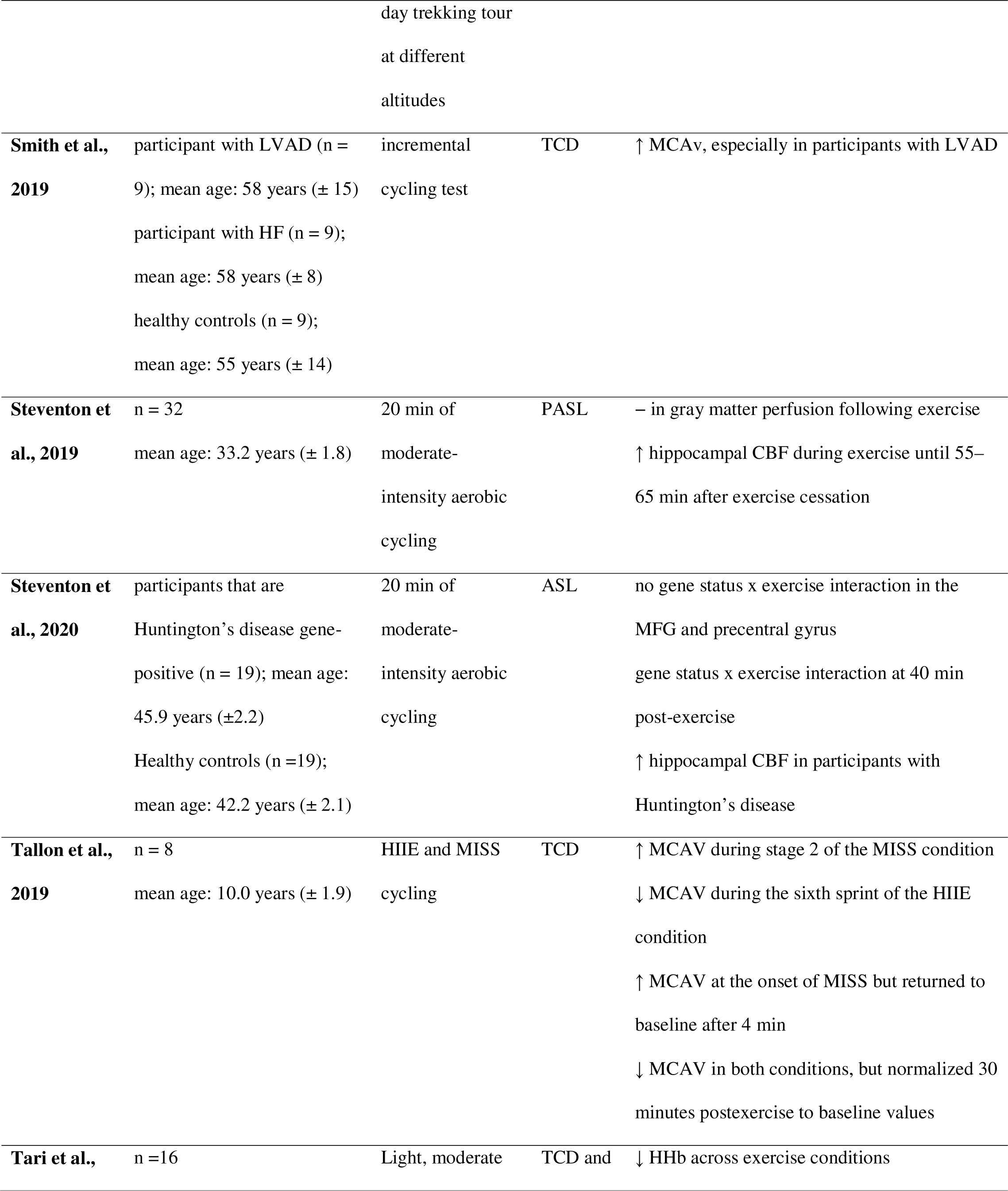
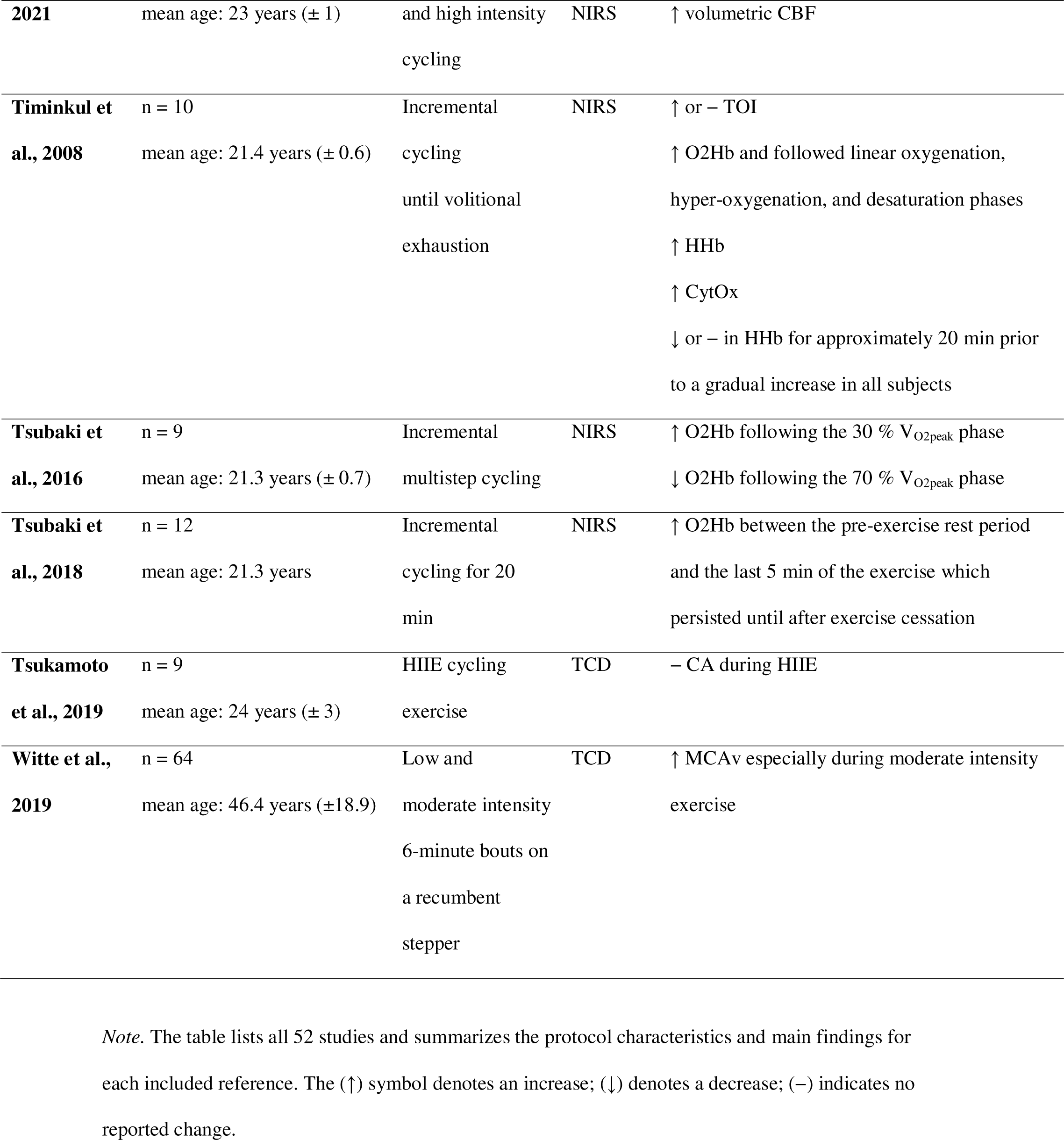
Study-level Results

| Study ID | Population | Intervention | Measure | Main Findings |
| --- | --- | --- | --- | --- |
| **Andrianopoulos et al., 2018** | patients with COPD (n= 52)<br>mean age: 68.2 years (± 9.0) | symptom-limited<br>CET at 75% of<br>the estimated<br>WR<sub>peak</sub> | NIRS | total-Hb ↑ in both CN and CI patients |
| **Ashley et al., 2020** | n = 26<br>mean age: 24 years (± 3.5) | GXT on a<br>recumbent cycle<br>ergometer | TCD | CBF response differences noted between sexes |
| **Billaut et al., 2010** | n = 11<br>mean age: 24.8 years (± 4.2)<br>trained | 5-km running<br>time trial on a<br>treadmill | fNIRS | ↑ Regional blood volume and Cox until 2.5 km<br>– between 2.5 km – 4.5 km<br>↓ 4.5 km -5 km |
| **Billinger et al., 2017** | n= 8 (low CV risk); mean<br>age: 23.88 years (± 1)<br>n=3 (high CV risk); mean<br>age: 66 years (± 1)<br>n = 1 (post ischemic stroke);<br>age: 65 years | incremental<br>recumbent stepper | TCD | ↑ MCAV (p=0.21) except for the individual<br>who had a history of stroke |
| **Billinger et al., 2021** | n = 22 (at 3- and 6-months<br>post-stroke)<br>mean age: 63.0 years (±14.0) | moderate intensity<br>recumbent stepper<br>exercise | TCD | – Cerebral blood velocity from 3- to 6-months<br>poststroke at rest and during an acute bout of<br>exercise |
| **Braz et al., 2017** | trained n = 20 (young: n=12;<br>older: n= 8)<br>untrained n = 20 (young: n<br>=11; older: n= 9)<br>mean age: young trained 23 | maximal<br>incremental on a<br>cycle ergometer | TCD | MCA V<sub>mean</sub> was ↓ with age<br>Bilateral ICA blood flow 31% higher in trained<br>compared to untrained individuals |
| | years ($\pm 4$); young untrained | | | |
| | 25 years ($\pm 5$); old trained 65 | | | |
| | years ($\pm 3$); old untrained 67 | | | |
| | years ($\pm 7$) | | | |
| **Burma et al., 2020** | n = 9<br>mean age: 26 years ($\pm 5$) | MICT and HIIT<br>performed on a<br>cycle ergometer | TCD | ↑ MCA during MICT<br>– CBV in the MCA and PCA during exercise<br>CA is impaired following exercise (6 h)<br>especially with HIIT |
| **Ellis et al., 2017** | n= 32<br>mean age: 9 years ($\pm 1$); n<br>=16<br>mean age: 25 years ($\pm 4$); n<br>=16 | maximal test on a<br>cycle ergometer | TCD | MCAV<sub>mean</sub> by age effect<br>MCAV<sub>mean</sub> by exercise intensity effect<br>↑ MCAV<sub>mean</sub> throughout exercise but lower for<br>children than adults |
| **Faulkner et al., 2016** | n=17<br>mean age: 24.6 years ($\pm 4.3$)<br>active | 1) GXTs to<br>maximal aerobic<br>capacity<br>2) Moderate load<br>submaximal bouts | NIRS | ↑ mean rSO<sub>2</sub> during recumbent compared with<br>upright exercise<br>↑ rSO<sub>2</sub> during moderate exercise |
| **Fisher et al., 2008** | n= 9 (young); mean age: 24<br>years ($\pm 3$)<br>n= 10 (middle-aged); mean<br>age: 57 years ($\pm 7$)<br>active | incremental<br>cycling | TCD | ↑ MCAV<sub>mean</sub> during low- and moderate-<br>intensity cycling ($P < 0.05$)<br>No CBF by age effect |
| **Fisher et al., 2013** | n = 11 (young; mean age: 22<br>years ($\pm 1$))<br>n = 9 (older; mean age: 66<br>years ($\pm 2$)) | incremental<br>maximal cycling | TCD | ↑ MCA V<sub>mean</sub> from rest to sub-maximal exercise<br>CBF by age effect |
|  | active |  |  |  |
| **Furlong et al., 2020** | n = 26<br>mean age: 26 years ($\pm 8$)<br>active | Incremental<br>running and<br>cycling | TCD | ↑ Mean MCAv from rest for both cycling and running |
| **Giles et al., 2014** | n = 24<br>mean age: 20.21 years ($\pm 2.38$)<br>active | 30 min cycling<br>exercise | fNIRS | ↑ O2Hb as a function of exercise load and duration<br>↑ dHb as a function of exercise load and duration<br>↑ tHb as a function of exercise load<br>↑ tHb level in the PFC as a function of exercise load and duration |
| **Hartmann et al., 2020** | n = 54 (smokers n = 27)<br>young; mean age group: 22 years ($\pm 1.6$)<br>middle-aged; mean age: 33 years ($\pm 7.8$) (smokers) and 36 years ($\pm 6.6$) (non-smokers)<br>active | 40 min constant<br>cycling at 50% of<br>$\text{VO}_{2\text{peak}}$ | NIRS | ↑ cerebral oxygenation in nonsmokers than smokers during exercise<br>exhibit |
| **Hiura et al., 2009** | n = 14<br>mean age = 20.5 years ($\pm 0.9$) | incremental<br>cycling until<br>volitional<br>exhaustion | NIRS | ↑ cerebral oxygenation with exercise |
| **Hiura et al., 2013** | n = 10<br>mean age: 22.7 years ($\pm 1.9$) | cycle ergometer | PET | ↑ rCBF during dynamic exercise |
| **Hiura et al., 2018** | n = 11<br>mean age: 22.5 years ($\pm 2.9$) | 20-min bout of<br>cycling (supine) | PET | ↑ rCBF at exercise in the midbrain, pons, medulla, cerebellar hemispheres and vermis, |
|  |  | cycle ergometer) |  | sensorimotor cortex for the bilateral legs<br>(M1Leg and S1Leg), right caudal nucleus,<br>right and left parietal operculum, insular<br>cortex, and right anterior and posterior<br>cingulate gyrus<br>↓ rCBF at post-10 exercise in the cerebellum,<br>inferior and superior temporal gyrus, left<br>thalamus, pons and left piriform lobe<br>↑ gCBF |
| **Ichinose et al., 2020** | n = 18 | intermittent bouts | NIRS | ↑ O2Hb of the left and right PFCs post-exercise (p < 0.01)<br>↑ HHb of the left and right PFCs post-exercise |
| **Imhoff et al., 2017** | n = 1<br>age: 35 years<br>trained | stepwise<br>incremental<br>upright cycling | TCD and<br>NIRS | ↑ MCAv<sub>mean</sub> with exercise |
| **Imray et al., 2005** | n = 11<br>mean age: 48.5 years (± 24.25) | Alticycle cycle<br>ergometer at<br>different altitudes | TCD and<br>NIRS | ↑ MCA blood velocity during exercise at 5,260<br>masl<br>– CVRest between the different altitudes<br>↓ cerebral oxygenation and oxygen delivery<br>during submaximal and maximal exercise at<br>altitude |
| **Klein et al., 2019** | young (mean age: 25 years (± 3); n = 11)<br>older (mean age: 69 years (± 3); n = 10) | submaximal<br>incremental<br>cycling | TCD | MCAv lower in the old than young<br>↑ MCAv during continuous exercise in the<br>young |
| **Koike et al., 2004a** | IDC patients, n = 35; mean<br>age: 57 years (±12) | incremental<br>symptom-limited | NIRS | ↑ O2Hb in healthy subjects during exercise<br>↓ O2Hb in 43% of the patients with IDC |
| | healthy control, n = 22; mean age: 56 years ($\pm 13$) | maximal cycling | | – O<sub>2</sub>Hb during 6 minutes of recovery especially in healthy subjects |
| **Koike et al., 2004b** | patients with VHD, n = 33<br>mean age: 64.7 years ($\pm 12.2$)<br>healthy control, n = 33 | Incremental cycling exercise | NIRS | ↑ O<sub>2</sub>Hb in healthy subjects during exercise<br>– dHb in healthy subjects during exercise<br>↓ O<sub>2</sub>Hb in patients with VHD during exercise<br>↑ dHb in patients with VHD during exercise |
| **Koike et al., 2006** | patients with IDC, n = 42;<br>mean age: 57 years ($\pm 12$)<br>healthy control, n = 29; mean age: 56 years ($\pm 14$) | incremental symptom-limited maximal cycling exercise | NIRS | ↑ O<sub>2</sub>Hb during exercise in 25 of 29 healthy subjects<br>↓ O<sub>2</sub>Hb during exercise in 18 of 42 patients with IDC |
| **Labrecque et al., 2020** | n = 10<br>mean age: 27 years ($\pm 6$)<br>trained | 30-s high-intensity cycling exercise | TCD | ↑ MCAv<sub>mean</sub> by 19 % ($\pm 10\%$) during exercise<br>↑ PCAv<sub>mean</sub> by 21 % ($\pm 14\%$) during exercise |
| **Lyngeraa et al., 2013** | n = 15<br>mean age: 31 years ($\pm 6.7$)<br>active | Incremental running bouts | TCD | ↑ MCA velocity with the onset of running to 65% and ↓ at 75%<br>↑ rCBF during exercise |
| **MacIntosh et al., 2017** | participants with BD, n = 31;<br>mean age: 16.9 years ($\pm 1.4$)<br>control subjects, n = 20;<br>mean age: 16.2 years ($\pm 1.7$) | single bout of cycling exercise | ASL | ↑ CBF at baseline in participants with BD in the left medial frontal gyrus and bilateral middle cingulate regions<br>↓ CBF 15 min post – exercise in participants with BD in the left precentral gyrus, left superior frontal gyrus, left occipital pole, right precentral gyrus, left postcentral gyrus, and the left frontal pole<br>larger ΔCBF ↓ in participants with BD compared to controls |
| **Malik et al.,** | n = 16 | HIIT | NIRS | ↑ ΔO<sub>2</sub>Hb from warm-up at work intervals 5 to |
| **2018** | mean age: 12 years ($\pm 1$) | | | 8<br>↑ ΔHHb from warm-up to work interval 8 during HIIE |
| **Matsukawa et al., 2015** | n = 22<br>mean age: 28 years ($\pm 2$) | cycling exercise for 30 s at 50 rpm | NIRS | ↑ prefrontal O<sub>2</sub>Hb following the exercise onset (peak at 5s with cued start)<br>↓ prefrontal O<sub>2</sub>Hb during later period of exercise and post-exercise period<br>– prefrontal dHb throughout exercise<br>↑ prefrontal dHb post-exercise |
| **Monroe et al., 2016** | n = 15<br>mean age: 21.3 years ($\pm 2.4$)<br>active | sprint and constant cycling sessions | NIRS | ↑ O<sub>2</sub>Hb during sprint<br>↑ dHb during sprint |
| **Neary et al., 2008** | CFS (n = 6); mean age: 39 years ($\pm 13$)<br>Control (n = 8); and mean age: 27 years ($\pm 7$) | maximal incremental cycling exercise | NIRS | ↑ HbO<sub>2</sub> during exercise until approximately 90% of time to exhaustion before plateauing or levelling off in both groups<br>↑ tHb during exercise until approximately 90% of time to exhaustion before plateauing or levelling off in both groups<br>↑ HHb during exercise until exercise termination in both groups<br>↓ TOI in both groups but more in the control group<br>Reduced cerebral oxygenation in participants with CFS compared to controls |
| **Parfitt et al., 2017** | n = 11<br>mean age: 27 years ($\pm 5$) | Treadmill 15-25 min bout either on land or in water | TCD | ↑ MCAv<sub>mean</sub> during exercise in both groups |
| **Perdomo et al., 2019** | n = 15<br>mean age: 45.5 years (± 8.9) | 10-minute running exercise bout (EX10) and 30-min running exercise bout (EX30) | TCD | ↑ mean MCA in both exercise bouts<br>↑ CBFv in both exercise bouts |
| **Pontifex et al., 2018** | n = 41<br>mean age: 10.2 years (± 1.0) | 20 min running exercise | ASL | ↓ cerebral blood flow in the left inferior frontal gyrus, extending across lateral prefrontal cortex, superior temporal gyrus, planum temporale, insular cortex, angular gyrus, inferior temporal cortex, and clusters in the anterior cingulate cortex and bilateral insular cortices |
| **Pugh et al., 2015** | n = 15<br>mean age: 26 years (± 4)<br>active | 20-min bouts of low-intensity stepping exercise either land or water based | TCD | ↑ MCAv associated with water-based exercise<br>↑ MCAv throughout both exercise protocols and returned to baseline after exercise<br>↑ PCAv throughout both exercise protocols and returned to baseline after exercise |
| **Rattray et al., 2017** | n = 15<br>mean age: 29 years (±14.5)<br>trained | 4 km time cycling trial | TCD | ↑ mean MCAv during exercise |
| **Robertson et al., 2015** | stroke supervisor (n= 13)<br>mean age: 64 years (± 15) | low- and moderate intensity cycling | ASL | ↓ CBF post-30 and post-50 in the right lentiform nucleus and left superior and middle temporal gyri<br>↑ CBF following moderate-intensity exercise in three clusters<br>↓ or – CBF following low-intensity exercise |
|  |  |  |  | ↑ in white matter CBF (33 %)<br>↓ rCBF following low-intensity exercise<br>↑ rCBF following moderate-intensity exercise<br>↓ CBF in the lentiform nucleus following up to 50 min both low- and moderate-intensity exercise |
| **Rupp & Perrey, 2008** | n = 13<br>trained<br>mean age: 24.9 years ($\pm 1.5$) | Progressive<br>maximal cycling<br>to exhaustion | NIRS | ↑ HbTot<br>↑ HHb<br>↑ HbO<sub>2</sub><br>↓ Cox following the first increase, before exhaustion |
| **Santos-Concejero et al., 2017** | n = 15<br>mean age: 23.7 years ($\pm 4.2$)<br>trained | 5-km running<br>time trial | NIRS | ↑ $\Delta$[O<sub>2</sub>Hb] during exercise<br>↓ $\Delta$[O<sub>2</sub>Hb] at the end of the trial<br>↓ $\Delta$[O<sub>2</sub>Hb] during the 30 s recovery periods<br>↑ $\Delta$[HHb] during exercise<br>↓ TOI |
| **Sato et al., 2011** | n = 10<br>mean age: 24 years ($\pm 5$) | Cycling | TCD | ↑ VA blood flow from rest to 80% V<sub>O<sub>2</sub>peak</sub><br>↑ CCA blood flow from rest to 80% V<sub>O<sub>2</sub>peak</sub><br>↑ MCA V<sub>mean</sub> from rest to 80% V<sub>O<sub>2</sub>peak</sub><br>↓ MCA V<sub>mean</sub> from 80% V<sub>O<sub>2</sub>peak</sub> |
| **Shibuya et al., 2004** | n = 6<br>mean age: 27.2 years ($\pm 0.6$) | intermittent<br>cycling | NIRS | ↑ Hb at the last two sets of intermittent exercise<br>↓ HbO<sub>2</sub> during the 15 s of recovery<br>↓ Hbdiff during the 15 s of recovery |
| **Smith et al., 2014** | n = 8<br>mean age: 30 years ($\pm 6$) | incremental<br>maximal cycling<br>test after a nine- | TCD | ↑ gCBF during exercise and recovery at high altitude (HA) compared with sea level (SL) |

|  |  |  |  |  |  |
| --- | --- | --- | --- | --- | --- |
|  |  | day trekking tour<br>at different<br>altitudes |  |  |  |
| **Smith et al., 2019** | participant with LVAD (n = 9); mean age: 58 years (± 15)<br>participant with HF (n = 9);<br>mean age: 58 years (± 8)<br>healthy controls (n = 9);<br>mean age: 55 years (± 14) | incremental<br>cycling test | TCD | ↑ MCAv, especially in participants with LVAD | |
| **Steventon et al., 2019** | n = 32<br>mean age: 33.2 years (± 1.8) | 20 min of<br>moderate-intensity aerobic<br>cycling | PASL | – in gray matter perfusion following exercise<br>↑ hippocampal CBF during exercise until 55–<br>65 min after exercise cessation | |
| **Steventon et al., 2020** | participants that are<br>Huntington's disease gene-positive (n = 19); mean age:<br>45.9 years (±2.2)<br>Healthy controls (n =19);<br>mean age: 42.2 years (± 2.1) | 20 min of<br>moderate-intensity aerobic<br>cycling | ASL | no gene status x exercise interaction in the<br>MFG and precentral gyrus<br>gene status x exercise interaction at 40 min<br>post-exercise<br>↑ hippocampal CBF in participants with<br>Huntington's disease | |
| **Tallon et al., 2019** | n = 8<br>mean age: 10.0 years (± 1.9) | HIIE and MISS<br>cycling | TCD | ↑ MCAV during stage 2 of the MISS condition<br>↓ MCAV during the sixth sprint of the HIIE<br>condition<br>↑ MCAV at the onset of MISS but returned to<br>baseline after 4 min<br>↓ MCAV in both conditions, but normalized 30<br>minutes postexercise to baseline values | |
| **Tari et al.,** | n =16 | Light, moderate | TCD and | ↓ HHb across exercise conditions | |
| **2021** | mean age: 23 years ($\pm 1$) | and high intensity cycling | NIRS | ↑ volumetric CBF |
| **Timinkul et al., 2008** | n = 10<br>mean age: 21.4 years ($\pm 0.6$) | Incremental cycling until volitional exhaustion | NIRS | ↑ or – TOI<br>↑ O2Hb and followed linear oxygenation, hyper-oxygenation, and desaturation phases<br>↑ HHb<br>↑ CytOx<br>↓ or – in HHb for approximately 20 min prior to a gradual increase in all subjects |
| **Tsubaki et al., 2016** | n = 9<br>mean age: 21.3 years ($\pm 0.7$) | Incremental multistep cycling | NIRS | ↑ O2Hb following the 30 % $V_{O2peak}$ phase<br>↓ O2Hb following the 70 % $V_{O2peak}$ phase |
| **Tsubaki et al., 2018** | n = 12<br>mean age: 21.3 years | Incremental cycling for 20 min | NIRS | ↑ O2Hb between the pre-exercise rest period and the last 5 min of the exercise which persisted until after exercise cessation |
| **Tsukamoto et al., 2019** | n = 9<br>mean age: 24 years ($\pm 3$) | HIIE cycling exercise | TCD | – CA during HIIE |
| **Witte et al., 2019** | n = 64<br>mean age: 46.4 years ($\pm 18.9$) | Low and moderate intensity 6-minute bouts on a recumbent stepper | TCD | ↑ MCAv especially during moderate intensity exercise |

*Note.* The table lists all 52 studies and summarizes the protocol characteristics and main findings for each included reference. The ( ↑ ) symbol denotes an increase; ( ↓ ) denotes a decrease; (–) indicates no reported change.

## 4. Discussion

This systematic review aimed to investigate the effect of cardiovascular exercise on cerebral perfusion in humans across a range of characteristics and modalities. Overall, exercise-induced changes in CBF appear to follow an inverted U- pattern, whereby CBF tends to increase at the beginning of an acute exercise intervention, before plateauing and returning to baseline levels toward the end of the bout or after exercise cessation. A potential reason for CBF following an inverted U-pattern is the prevention of hypoperfusion and CA maintenance. Exercise-induced increases in CBF allow for an increase in the supply of oxygen and other nutrients to the brain to cope with the heightened demands associated with exercise. However, if CBF would continue to increase across the entire duration of an exercise intervention, the risk for hypoperfusion might significantly increase. Therefore, the brain seems to have mechanisms in place that allow for CBF to return to baseline levels towards the end of an exercise intervention or post-exercise. Beyond exercise duration, exercise intensity appears to play an important role in exercise-induced changes in CBF.

Across the 52 included studies, the myriad of impacting factors that may shape the effect acute cardiovascular exercise has on cerebral perfusion were particularly salient. Among those potential mediating factors were sex, age, altitude, health history, and fitness level in the realm of participant characteristics. Several additional factors were identified, including exercise intensity, exercise duration, and the technique used to measure CBF. The following subsections detail the risk of bias assessment performed on each of the 52 included studies, the study characteristics, including interventions and CBF measurement techniques used, and some overall limitations that we identified.

### 4.1. Risk of bias

Overall, the included studies were thought to have a low risk of bias. However, the domain of randomization presented a more significant challenge for 42 out of 52 papers compared to other domains. A potential reason for 80.77% of the included studies getting a ”some concern” rating in the domain of randomization might be procedures not being described in sufficient detail. Some studies may not report their procedures in sufficient detail, while others do not mention blinding and randomization processes because they were not part of the experiment. Notably, the blinding of participants may be practically difficult in some exercise experiments because participants notice whether they are exercising or not and, therefore, in what group they are likely to be. However, strategies such as asking all participants to exercise but at different intensities (e.g., Pontifex et al., 2018), incorporating other conditions such as sleep to reduce expectation biases and other confounding variables such as posture may lower the overall risk of bias and therefore, increase the quality of a study.

### 4.2. Key Participant Characteristics Modulating CBF

The present review provides insight into the impact of sex, age, altitude, health history, and fitness level on the effect of acute cardiovascular exercise on CBF. First, sex seems to significantly shape the effect acute cardiovascular exercise has on cerebral perfusion in humans. 61.93 % of the combined sample included in the present review identified as male. However, few studies investigated potential sex differences. However, one study that did thematize sex as a moderator in the acute exercise CBF relationship reports crucial differences between the sexes. Ashley and colleagues (2020) found a significantly higher CPP in males than females. Simultaneously, females showed more significant dilation in the cerebral vasculature (i.e., conductance, ΔCVCi) in the periphery than males. As exercise intensity increased, males showed a more substantial reduction in ΔCVCi compared to females at 100%Wmax. Ashley and colleagues (2020) hypothesize that hormone concentrations, sympathetic outflow, and flow-mediated dilation may be among the mechanisms that could drive sex differences in the CBF response to acute exercise. However, more research in this area is needed to better understand the modulating effect of sex within the relationship between acute exercise and CBF, and specifically the precise mechanisms that drive presumed sex differences.

Second, age seems to play an important role in acute cardiovascular exercise’s effect on CBF. Included studies that looked at the effect of age on CBF in the realm of exercise underline a clear trend: age negatively correlates with cerebral perfusion. Older participants tend to show lower CBF than younger participants (Braz et al., 2017; Ellis et al., 2017; Fisher et al., 2013; Klein et al., 2019). However, in children, CBF in the anterior cerebral circulation is approximately half as high compared to that of adults (Ellis et al., 2017). The age group represented the most in the current systematic review is young adults aged between 18 and 29 years. Young adults (18-29 years) being the most common age group participating in the included papers seems to mirror a trend in psychological research to rely on student samples due to lower administration costs, assumed lower response bias, and perceived facility of recruitment (Hanel & Vione, 2016). Student samples may not accurately reflect the population, and thus, findings may not be generalizable to the general public (Hanel & Vione, 2016). However, in the context of the present topic, students may accurately represent non-students with similar age, sex, health, and fitness histories.

Third, the altitude at which exercise is carried out could play a critical role in the effect cardiovascular exercise has on cerebral perfusion and the recovery period. Smith and colleagues (2014) found that acute cardiovascular exercise differentially affects cerebral perfusion depending on whether individuals exercise at high altitudes (>3500 masl) or sea level. GCBF has been associated with increasing substantially more during exercise and recovery at high altitudes. Smith and colleagues (2014) interpreted the more significant increase in gCBF during exercise at high altitudes as a compensatory mechanism to maintain cerebral oxygen delivery. Akin to earlier potential moderators in the acute exercise- CBF relationship, the effects of altitude on CBF during exercise call for further research.

Fourth, health history can shape the effect of acute cardiovascular exercise on CBF depending on the relevance of a given condition. Conditions such as IDC, COPD, VHD, BD, stroke, Huntington’s Disease, heart failure, and CFS that predominantly affect the heart, lungs, and brain play significant roles in the exercise- CBF relationship. For instance, BD has been associated with increased CBF in the left medial frontal gyrus and bilateral middle cingulate regions at baseline and greater decreases in CBF fifteen minutes after exercise cessation (MacIntosh et al., 2017). MacIntosh and colleagues (2017) explain the elevated CBF in adolescents with BD by drawing attention to potential delays in frontal lobe maturation that could drive elevations in CBF. Further studies looking at the role BD may play in the effect acute cardiovascular exercise may have on the CBF of adults with BD are needed to derive a complete picture of the role of BD in CBF and exercise. Similarly, CFS has been linked with reduced cerebral oxygenation (Neary et al., 2008). Neary and colleagues (2008) argued the reduced cerebral oxygenation and blood volume changes were associated with the reduction in regional cerebral blood flow that typically manifest with CFS. However, similar to the aforementioned study, Neary and colleagues (2008) also call for further studies investigating the CFS-CBF-exercise relationship.

Finally, fitness level has been shown to play a critical role in the effect acute cardiovascular exercise has on cerebral oxygenation. Trained and active people may be in an advantageous position in terms of potential exercise-induced effects on CBF compared to sedentary people. For instance, Braz and colleagues (2017) report that the bilateral ICA blood flow is 31% higher in trained compared to untrained participants. This could lead to an increase in nutrient supply to the brain and aid brain function, including cognition. Habits such as smoking, on the other hand, may limit the increase in gCBF associated with acute exercise (Hartmann et al., 2020).

### 4.3. Exercise Characteristics

An acute bout of exercise can be characterized by exercise type, exercise intensity, and exercise duration. First, the exercise types included in the present review range from cycling to running, performing stepping exercises, engaging in other forms of HIIT, or combinations of the aforementioned exercise modalities. Overall, cycling was the most common exercise intervention employed, with 76.9 % of all 52 studies using electronically braked ergometer cycling. 13.5 % of studies used running on either land- or water-based treadmills for the exercise intervention. Exercise on recumbent steppers was the third most common exercise modality, with 7.7 % using it. Lastly, 3.8 % of included studies employed a form of HIIT that does not include cycling, running on a treadmill, or stepping. In their study, Furlong and colleagues (2020) decided to use both cycling on an electronically braked cycle ergometer and treadmill running.

Second, exercise intensity can either vary or stay constant across the exercise protocol. In the present review, the various exercise protocols were classified as incremental exercise (59.6 % of all 52 studies), constant (40.4 %), interval (21.2 %), or time trial interventions (3.8 %). However, 25 % of all studies employed mixed exercise interventions where participants completed, for example, one incremental and one constant running trial. Furthermore, the actual exercise intensity employed either in a constant or a graded exercise protocol can be measured in various ways. Exercise intensity can be specified and operationalized using either parameters of internal or external load (Herold et al., 2020). For example, defining exercise intensity through Vo2max can be considered using parameters of internal loads. On the other hand, using the watt output on a cycle ergometer as a surrogate measure of exercise intensity is an example of employing parameters of external load. Included studies using parameters of internal load to determine exercise intensity (Andrianopoulos et al., 2018; Billaut et al., 2010; Billinger et al., 2017; Billinger et al., 2021; Burma et al., 2020; Ellis et al., 2017; Faulkner et al., 2016; Fisher et al., 2013; Furlong et al., 2020; Giles et al., 2014; Hiura et al., 2009; Hiura et al., 2013; Hiura et al., 2018; Ichinose et al., 2020; Klein et al., 2019; Labrecque et al., 2020; Lyngeraa et al., 2013; Malik et al., 2018; Monroe et al., 2016; Parfitt et al., 2017; Perdomo et al., 2019; Pontifex et al., 2018; Rattray et al., 2017; Robertson et al., 2015; Santos-Concejero et al., 2017; Sato et al., 2011; Shibuya et al., 2004; Steventon et al., 2019; Steventon et al., 2020; Tsubaki et al., 2018; Tsubaki et al., 2016; Witte et al., 2019) focused on Vo2max, heart rate reserve (HRR), peak work rate (WRpeak), peak oxygen uptake (VOpeak), and perceived exertion rates (Borg, 1970). On the other hand, experiments using parameters of external load centered around Watt (W) as a measure of exercise intensity (Ashley et al., 2020; Billinger et al., 2017; Braz et al., 2017; Fisher et al., 2013; Fisher et al., 2008; Hartmann et al., 2020; Hiura et al., 2009; Hiura et al., 2018; Imhoff et al., 2017; Imray et al., 2005; Koike et al., 2004a; Koike et al., 2004b; Koike et al., 2006; Neary et al., 2008; Rupp & Perrey, 2008; Smith et al., 2019; Tallon et al., 2019; Tari et al., 2021; Timinkul et al., 2008; Tsukamoto et al., 2019). The internal load is influenced by personal characteristics, including nutrition, genetics, sleep, fitness, health history, and similar, and environmental factors such as room temperature. Several studies aimed at keeping room temperature constant across individuals (e.g., Billaut et al., 2010; Billinger et al., 2017; Billinger et al., 2021). For instance, Billaut and colleagues (2010) kept room temperature in the laboratory between 18 °C and 20 °C, while Billinger and colleagues (2017; 2021) chose temperatures between 22°C and 24°C. Burma and colleagues (2020) controlled the diet participants consumed across the day by providing meal replacement shakes, and Santos-Concejero and colleagues (2017) controlled for altitude (130 masl). Moreover, exercise intensities were reported inconsistently across the studies included in the present review. Various labels were employed ranging from maximal, submaximal, high, moderate, low to using Borg scale ratings of perceived exertion (Borg, 1970). The Borg category-ratio scale (Borg, 1970) ranges from 6 to 20, with 11 being “fairly light” (p. 93), 15 being “hard” (p. 93), and 19 standing for “very, very hard” (p. 93). For example, Billaut and colleagues (2010) asked participants to reflect on their perceived exertion and give a Borg scale rating every 0.5 km during a 5km running time trial. The ratings increased from 6.6 ± 0.6 at baseline to 12.9 ± 1.7 at 0.5 km and 19.1 ± 0.7 at 5km (Billaut et al., 2010). In other words, the participants went from perceiving the treadmill running as “very, very light” (Borg, 1970, p. 93) at baseline to rating it “very, very hard” (Borg, 1970, p. 93) at the end of the 5km run. This perceived exertion scale may be useful to capture participants’ perceptions of exercise intensity and their fitness, but it cannot be equated to other labels that are based on VO2max. Different studies employed different definitions of “submaximal” exercise. For example, Klein and colleagues (2019) defined “submaximal” exercise as participants reaching a heart rate that corresponds to 85% of their age-predicted maximum (220 - age). On the other hand, Faulkner and colleagues (2016) describe submaximal exercise as participants reaching 45% to 60% VO2max. Furthermore, 32.7 % of all included studies used maximal exercise, while 48.1 % of studies aimed for high intensities (≥ 70% HRR; Andrianopoulos et al., 2018), 40.4 % used moderate-intensity exercise (∼45- 70 % HRR; Hiura et al., 2018; Robertson et al., 2015), and 17.3 % employed low exercise intensity (≤ 44 % HRR; Hiura et al., 2018; Robertson et al., 2015). 38.5 % of included studies included several exercise intensities. However, using Vo2max-based categories to describe exercise intensities may have some limitations (Warpeha, 2003). First, although there seems to be an upper limit to oxygen uptake for each person, this limit seems to vary significantly across individuals (Warpeha, 2003). Pulmonary, peripheral limitations to oxygen extraction at the cellular level, and cardiac factors influence Vo2max (Warpeha, 2003). This might explain the different definitions of what, for instance, “submaximal” exercise means. Overall, exercise intensities were reported. Third, exercise durations used in the included studies ranged from 6 (Billinger et al., 2021; Tallon et al., 2019) to 45 minutes (Burma et al., 2020). Overall, exercise durations can be grouped into three categories: ≤ 15 minutes (Billinger et al., 2021; Hiura et al., 2013; Imray et al., 2005; Klein et al., 2019; Labrecque et al., 2020; Lyngeraa et al., 2013; Malik et al., 2018; Neary et al., 2008; Sato et al., 2011; Tallon et al., 2019; Tari et al., 2021; Witte et al., 2019), between 16 minutes and 30 minutes (Billaut et al., 2010; Burma et al., 2020; Faulkner et al., 2016; Fisher et al., 2008; Furlong et al., 2020; Giles et al., 2014; Hiura et al., 2018; Ichinose et al., 2020; MacIntosh et al., 2017; Monroe et al., 2016; Parfitt et al., 2017; Perdomo et al., 2019; Pontifex et al., 2018; Pugh et al., 2015; Robertson et al., 2015; Steventon et al., 2019; Steventon et al., 2020; Tsubaki et al., 2018; Tsubaki et al., 2016), and ≥ 31 minutes (Burma et al., 2020; Hartmann et al., 2020; Rattray et al., 2017). Several studies did not report clear exercise duration, while others employed several different exercise durations due to the use of several exercise protocols. The product of exercise duration, intensity, and type is referred to as the dose of activity (Pontifex et al., 2018). However, the dose-response relationship between acute aerobic exercise or HIIT and CBF remains poorly understood. This is partly due to inadequate exercise description and heterogeneous measures of exercise intensity.

### 4.4. Cerebral Blood Flow Measurement Characteristics

The included studies chose to measure CBF via TCD (n =25), NIRS (n = 23), ASL (n = 5), PET (n = 2), or combinations of those techniques (n = 3). All of those four imaging techniques come with advantages and limitations. TCD does not directly measure CBF (Lipnick et al., 2018). Instead, TCD measures blood flow velocity as it does not consider vessel diameter (Foster et al., 2019). However, if the angle of insonation remains unchanged during imaging, changes in TCD velocities are likely to reflect changes in middle cerebral artery blood flow rate (Lipnick et al., 2018). Lipnick and colleagues (2018) highlight that collecting accurate TCD measures requires adequate transcranial windows and trained individuals to perform and interpret the TCD ultrasound. Also, continuous measurements may be difficult and subject to electrical noise interference (Lipnick et al., 2018). Nevertheless, TCD is non-invasive, less expensive than other imaging techniques (Lipnick et al., 2018), and offers a high temporal resolution. Akin to TCD, NIRS also offers high temporal resolution, is portable, and is non-invasive (Shibuya et al., 2004). However, like TCD, NIRS signals are not a direct measure of CBF (Lipnick et al., 2018). NIRS measures cerebral tissue oxygenation in the first few millimeters of the frontal cortex (Cardim & Griesdale, 2018) and depends on arteriovenous hemoglobin concentration and oxygenation (Lipnick et al., 2018). A potential limitation to using NIRS to measure CBF is the inability of this technique to evaluate more than a small region of the brain (Cardim & Griesdale, 2018; Lipnick et al., 2018). Also, NIRS is sensitive to extracranial blood flow and skin vasculature signals, which may contaminate regional cerebral oxygen saturation (rSO2) (Auger et al., 2016; Cardim & Griesdale, 2018; Matsukawa et al., 2015). Attempting to address the limitations of NIRS techniques, the ultrasound-tagged NIRS (UT-NIRS) has been developed (Cardim & Griesdale, 2018; Lipnick et al., 2018). The UT-NIRS uses low-power ultrasound to modulate NIR light (Cardim & Griesdale, 2018). The photons then undergo a Doppler effect within the targeted tissue which is interpreted as a result of blood cell movement (Cardim & Griesdale, 2018). The velocity of the blood cell movement serves as a surrogate measure of CBF, called the cerebral flow index (CFI) (Cardim & Griesdale, 2018).

Again, CBF has no fixed relationship with cerebral blood volume (CBV) or blood velocity (Buxton, 2005). Therefore, Buxton (2005) argues that the most robust quantitative measurement of CBF is quantifying the delivery of an agent carried in arterial blood. ASL techniques are non-invasive imaging applications where arterial blood gets tagged (the magnetization gets inverted), and after a delay, the tagged blood reaches the image plane where an image is acquired (Buxton, 2005). Then a second measurement is taken without tagging the blood. Subsequentially, the tag and control images are adjusted, and the difference signal (amount of tagged blood delivered to voxel at time of measurement) is proportional to CBF (Buxton, 2005). However, the transit delays from the tagging region to the image voxel may constitute a systematic error (Buxton, 2005). Buxton (2005) identifies three principal factors that need to be considered when using ASL to quantify CBF. First, producing an accurate control (without tagging the arterial blood) measurement is crucial (Buxton, 2005). Second, creating a well-defined tagged blood- bolus and carefully checking that all of the bolus is delivered to an imaging voxel is key (Buxton, 2005). Third, relaxation and potential clearance have to be considered here because of a potential reduction in the magnitude of the tagged magnetization (Buxton, 2005). Moreover, due to ASL being relatively expensive (Lipnick et al., 2018), sample size and replication tend to fall short. However, the included experiments in this systematic review that used ASL MRI had sample sizes of n = 51 (MacIntosh et al., 2017), n = 41 (Pontifex et al., 2018), n = 13 (Robertson et al., 2015), n = 32 (Steventon et al., 2019), and n = 38 (Steventon et al., 2020). Like ASL MRI, PET also relies on measuring contrast agents that blood flow delivered and cleared from cerebral tissue (Liu & Brown, 2007). In other words, PET necessitates the injection of radioactive contrast agents (Auger et al., 2016). Furthermore, like ASL, PET is not continuous and relatively cost-intensive (Lipnick et al., 2018).

Studies were difficult to compare due to the heterogeneity in CBF measures used and the variability across samples regarding health characteristics, anthropomorphic data, and exercise protocols. However, several studies report that the acute exercise-CBF relationship tends to follow an inverted-U response, whereby CBF tends to increase at the onset of exercise, before then plateauing and finally decreasing towards the peak of or at the end of the exercise. Similarly, whether CA remains stable in the event of acute cardiovascular exercise could not be answered by this review due to conflicting results reported in included studies. While Tsukamoto and colleagues (2019) argue for CA during HIIT to remain unaffected, Burma and colleagues (2020) report that CA was impaired following HIIT. Taken together, typically, CBF seems to increase during acute cardiovascular exercise, but several moderators, including age, sex, fitness, health history, altitude, and exercise intensity, seem to influence the effect a single bout of exercise can have on CBF.

## 6. Conclusion

Overall, the studies included in this systematic review were mixed, but clearly indicate a general trend for the acute cardiovascular exercise-CBF relationship to follow an inverted U- response. CBF typically increases at the onset of a single bout of exercise, to then plateau before decreasing at the end of a bout or after exercise ceases, most notably in the insular cortex, the midbrain, the right and left parietal operculum, the pons, the medulla, the right anterior and posterior cingulate gyrus, the right caudal nucleus, the sensorimotor cortex, the cerebellar hemispheres and vermis, and the hippocampus. Many factors appear to play an important role in this relationship, and several of them remain understudied. Beyond furthering knowledge about the human brain, a better understanding of the factors influencing exercise-induced change in CBF has the potential to help refine and personalize interventions that can promote brain health across a range of populations.

## Data Availability

All data produced in the present work are contained in the manuscript.

## References

1. Ainslie, P.N., Cotter, J.D., George, K.P., Lucas, S., Murrell. C, Shave, R., Thomas, K. N., Williams, M. J. A., & Atkinson, G. (2008). Elevation in cerebral blood flow velocity with aerobic fitness throughout healthy human ageing. The Journal of Physiology, 586(16), 4005–4010. https://doi.org/10.1113/jphysiol.2008.158279

2. Anazodo, U. C., Shoemaker, J. K., Suskin, N., Ssali, T., Wang, D. J. J., & St. Lawrence, K. S. (2016). Impaired cerebrovascular function in coronary artery disease patients and recovery following cardiac rehabilitation. Frontiers in Aging Neuroscience, 7. https://doi.org/10.3389/fnagi.2015.00224

3. Andrianopoulos, V., Vogiatzis, I., Gloeckl, R., Bals, R., Koczulla, R. A., & Kenn, K. (2018). Cerebral oxygen availability during exercise in COPD patients with cognitive impairment. Respiratory Physiology & Neurobiology, 254, 64–72. https://doi.org/10.1016/j.resp.2018.05.001

4. Ashley, J. D., Shelley, J. H., Sun, J., Song, J., Trent, J. A., Ambrosio, L. D., Larson, D. J., Larson, R. D., Yabluchanskiy, A., & Kellawan, J. M. (2020). Cerebrovascular responses to graded exercise in young healthy males and females. Physiological Reports, 8(20). https://doi.org/10.14814/phy2.14622

5. Auger, H., Bherer, L., Boucher, É., Hoge, R., Lesage, F., & Dehaes, M. (2016). Quantification of extra-cerebral and cerebral hemoglobin concentrations during physical exercise using time-domain near infrared spectroscopy. Biomedical Optics Express, 7(10), 3826. https://doi.org/10.1364/boe.7.003826

6. Bailey, D. M., Marley, C. J., Brugniaux, J. V., Hodson, D., New, K. J., Ogoh, S., & Ainslie, P. N. (2013). Elevated aerobic fitness sustained throughout the adult lifespan is associated with improved cerebral hemodynamics. Stroke, 44(11), 3235–3238. https://doi.org/10.1161/strokeaha.113.002589

7. Barnes, J. N. (2015). Exercise, cognitive function, and aging. Advances in Physiology Education, 39(2), 55–62. https://doi.org/10.1152/advan.00101.2014

8. Biagi, L., Abbruzzese, A., Bianchi, M. C., Alsop, D. C., Del Guerra, A., & Tosetti, M. (2007). Age dependence of cerebral perfusion assessed by magnetic resonance continuous arterial spin labeling. Journal of Magnetic Resonance Imaging, 25(4), 696–702. https://doi.org/10.1002/jmri.20839

9. Billaut, F., Davis, J. M., Smith, K. J., Marino, F. E., & Noakes, T. D. (2010). Cerebral oxygenation decreases but does not impair performance during self-paced, strenuous exercise. Acta Physiologica, 198(4), 477–486. https://doi.org/10.1111/j.1748-1716.2009.02058.x

10. Billinger, S. A., Craig, J. C., Kwapiszeski, S. J., Sisante, J. V., Vidoni, E. D., Maletsky, R., & Poole, D. C. (2017). Dynamics of middle cerebral artery blood flow velocity during moderate-intensity exercise. Journal of applied physiology, 122(5), 1125– 1133. https://doi.org/10.1152/japplphysiol.00995.2016

11. Billinger, S. A., Whitaker, A. A., Morton, A., Kaufman, C. S., Perdomo, S. J., Ward, J. L., Eickmeyer, S. M., Bai, S. X., Ledbetter, L., & Abraham, M. G. (2021). Pilot study to characterize middle cerebral artery dynamic response to an acute bout of moderate intensity exercise at 3L and 6Lmonths poststroke. Journal of the American Heart Association, 10(3), 10: e017821. https://doi.org/10.1161/JAHA.120.017821

12. Borg, G. (1970). Perceived exertion as an indicator of somatic stress. Scandinavian Journal of Rehabilitation Medicine, 2(2), 92–98. https://doi.org/10.2340/1650197719702239298

13. Braz, I. D., Flück, D., Lip, G. Y. H., Lundby, C., & Fisher, J. P. (2017). Impact of aerobic fitness on cerebral blood flow and cerebral vascular responsiveness to CO2 in young and older men. Scandinavian Journal of Medicine & Science in Sports, 27(6), 634–642. https://doi.org/10.1111/sms.12674

14. Burma, J. S., Copeland, P., Macaulay, A., Khatra, O., Wright, A. D., & Smirl, J. D. (2020). Dynamic cerebral autoregulation across the cardiac cycle during 8 hr of recovery from acute exercise. Physiological Reports, 8(5). https://doi.org/10.14814/phy2.14367

15. Buxton, R. B. (2005). Quantifying CBF with arterial spin labeling. Journal Of Magnetic Resonance Imaging, 22, 723–726. https://doi.org/10.1002/jmri.20462

16. Cardim, D., & Griesdale, D. E. (2018). Near-infrared spectroscopy: Unfulfilled promises. British Journal of Anaesthesia, 121(3), 523–526. https://doi.org/10.1016/j.bja.2018.05.058

17. Chapman, S. B., Aslan, S., Spence, J. S., DeFina, L. F., Keebler, M. W., Didehbani, N., & Lu, H. (2013). Shorter term aerobic exercise improves brain, cognition, and cardiovascular fitness in aging. Frontiers in Aging Neuroscience, 5. https://doi.org/10.3389/fnagi.2013.00075

18. Chen, W.-L., Wagner, J., Heugel, N., Sugar, J., Lee, Y.-W., Conant, L., Malloy, M., Heffernan, J., Quirk, B., Zinos, A., Beardsley, S. A., Prost, R., & Whelan, H. T. (2020). Functional near-infrared spectroscopy and its clinical application in the field of neuroscience: Advances and future directions. Frontiers in Neuroscience, 14. https://doi.org/10.3389/fnins.2020.00724

19. Clausen, M., Pendergast, D. R., Wilier, B., & Leddy, J. (2016). Cerebral blood flow during treadmill exercise is a marker of physiological postconcussion syndrome in female athletes. Journal of Head Trauma Rehabilitation, 31(3), 215–224. https://doi.org/10.1097/HTR.0000000000000145

20. Colcombe, S. J., Erickson, K. I., Scalf, P. E., Kim, J. S., Prakash, R., McAuley, E., Elavsky, S., Marquez, D. M., Hu, L., & Kramer, A. F. (2006). Aerobic exercise training increases brain volume in aging humans. The Journals of Gerontology Series A: Biological Sciences and Medical Sciences, 61(11), 1166–1170. https://doi.org/10.1093/gerona/61.11.1166

21. Colcombe, S., & Kramer, A. F. (2003) Fitness effects on the cognitive function of older adults: A meta-analytic study. Psychological Science, 14, 125–130. https://journals.sagepub.com/doi/pdf/10.1111/1467-9280.t01-1-01430?casa_token=bt1KOpdXIwsAAAAA%3AUrKGGemkzVIVuTZKx7x41WMNSqU6bdL4_rLq7g41f9rgIDSGOQD8d4jAQQdUItOlK2Rb_oYTwirzrhg&

22. Del Zoppo, G. J., Sharp, F. R., Heiss, W.-D., & Albers, G. W. (2011). Heterogeneity in the penumbra. Journal of Cerebral Blood Flow & Metabolism, 31(9), 1836–1851. https://doi.org/10.1038/jcbfm.2011.93

23. Ellis, L. A., Ainslie, P. N., Armstrong, V. A., Morris, L. E., Simair, R. G., Sletten, N. R., Tallon, C. M., & McManus, A. M. (2017). Anterior cerebral blood velocity and end-tidal CO2 responses to exercise differ in children and adults. American Journal of Physiology-Heart and Circulatory Physiology, 312(6), H1195–H1202. https://doi.org/10.1152/ajpheart.00034.2017

24. Faulkner, J., Lambrick, D., Kaufmann, S., & Stoner, L. (2016). Effects of upright and recumbent cycling on executive function and prefrontal cortex oxygenation in young healthy men. Journal of Physical Activity and Health, 13(8), 882–887. https://doi.org/10.1123/jpah.2015-0454

25. Fisher, J. P., Hartwich, D., Seifert, T., Olesen, N. D., McNulty, C. L., Nielsen, H. B., van Lieshout, J. J., & Secher, N. H. (2013). Cerebral perfusion, oxygenation and metabolism during exercise in young and elderly individuals. The Journal of physiology, 591(7), 1859–1870. https://doi.org/10.1113/jphysiol.2012.244905

26. Fisher, J. P., Ogoh, S., Young, C. N., Raven, P. B., & Fadel, P. J. (2008). Regulation of middle cerebral artery blood velocity during dynamic exercise in humans: Influence of aging. Journal of Applied Physiology, 105(1), 266–273. https://doi.org/10.1152/japplphysiol.00118.2008

27. Flück, D., Braz, I. D., Keiser, S., Hüppin, F., Haider, T., Hilty, M. P., Fisher, J. P., & Lundby, C. (2014). Age, aerobic fitness, and cerebral perfusion during exercise: Role of carbon dioxide. American Journal of Physiology-Heart and Circulatory Physiology, 307(4), H515–H523. https://doi.org/10.1152/ajpheart.00177.2014

28. Foster, C., Steventon, J. J., Helme, D., Tomassini, V., & Wise, R. G. (2019). Assessment of the effects of aerobic fitness on cerebrovascular function in young adults using multiple inversion time arterial spin labelling MRI. BioRxiv. https://doi.org/10.1101/539072

29. Furlong, R. J., Weaver, S. R., Sutherland, R., Burley, C. V., Imi, G. M., Lucas, R., & Lucas, S. (2020). Exercise-induced elevations in cerebral blood velocity are greater in running compared to cycling at higher intensities. Physiological reports, 8(15), e14539. https://doi.org/10.14814/phy2.14539

30. Giles, G. E., Brunyé, T. T., Eddy, M. D., Mahoney, C. R., Gagnon, S. A., Taylor, H. A., & Kanarek, R. B. (2014). Acute exercise increases oxygenated and deoxygenated hemoglobin in the prefrontal cortex. NeuroReport, 25(16), 1320–1325. https://doi.org/10.1097/wnr.0000000000000266

31. Hanel, P. H. P., & Vione, K. C. (2016). Do student samples provide an accurate estimate of the general public? PLOS ONE, 11(12), e0168354. https://doi.org/10.1371/journal.pone.0168354

32. Hartmann, T. E., Marino, F. E., & Duffield, R. (2020). The effect of cigarette smoking history on autonomic and cerebral oxygenation responses to an acute exercise bout in smokers. Physiological Reports, 8(19). http://dx.doi.org/10.14814/phy2.14596

33. Herold, F., Aye, N., Lehmann, N., Taubert, M., & Müller, N. G. (2020). The contribution of functional magnetic resonance imaging to the understanding of the effects of acute physical exercise on cognition. Brain Sciences, 10(3), 175, 1-32. https://doi.org/10.3390/brainsci10030175

34. Hiura, M., Mizuno, T., & Fujimoto, T. (2009). Cerebral oxygenation in the frontal lobe cortex during incremental exercise tests: The regional changes influenced by volitional exhaustion. Advances in Experimental Medicine and Biology, 662, 257–263. https://doi.org/10.1007/978-1-4419-1241-1_37

35. Hiura, M., Nariai, T., Ishii, K., Sakata, M., Oda, K., Toyohara, J., & Ishiwata, K. (2013). Changes in cerebral blood flow during steady-state cycling exercise: A study using oxygen-15-labeled water with PET. Journal of Cerebral Blood Flow & Metabolism, 34(3), 389–396. https://doi.org/10.1038/jcbfm.2013.220

36. Hiura, M., Nariai, T., Sakata, M., Muta, A., Ishibashi, K., Wagatsuma, K., Tetsuro, K., Jun Toyohara, T., Ishii, K., & Maehara, T. (2018). Response of cerebral blood flow and blood pressure to dynamic exercise: A study using PET. International Journal of Sports Medicine, 39(03), 181–188. https://doi.org/10.1055/s-0043-123647

37. Ichinose, Y., Morishita, S., Suzuki, R., Endo, G., & Tsubaki, A. (2020). Comparison of the effects of continuous and intermittent exercise on cerebral oxygenation and cognitive function. Advances in Experimental Medicine and Biology, 209–214. https://doi.org/10.1007/978-3-030-34461-0_26

38. Imhoff, S., Malenfant, S., Nadreau, É., Poirier, P., Bailey, D. M., & Brassard, P. (2017). Uncoupling between cerebral perfusion and oxygenation during incremental exercise in an athlete with postconcussion syndrome: A case report. Physiological Reports, 5(2), e13131. https://doi.org/10.14814/phy2.13131

39. Imray, C. H. E., Myers, S. D., Pattinson, K. T. S., Bradwell, A. R., Chan, C. W., Harris, S., Collins, P., Wright, A. D., & the Birmingham Medical Research Expeditionary Society (2005). Effect of exercise on cerebral perfusion in humans at high altitude. Journal of Applied Physiology, 99(2), 699–706. https://doi.org/10.1152/japplphysiol.00973.2004

40. Klein, T., Bailey, T., Abeln, V., Schneider, S., & Askew, C. (2019). Cerebral blood flow during interval and continuous exercise in young and old men. Medicine & Science in Sports & Exercise, 51, 1523–1531. https://doi.org/10.1249/MSS.0000000000001924

41. Koike, A., Hoshimoto, M., Nagayama, O., Tajima, A., Kubozono, T., Oikawa, K., Uejima, T., Momose, T., Aizawa, T., Fu, L.T., & Itoh, H. (2004a). Cerebral oxygenation during exercise and exercise recovery in patients with idiopathic dilated cardiomyopathy. The American Journal of Cardiology, 94(6), 821–824. https://doi.org/10.1016/j.amjcard.2004.06.013

42. Koike, A., Hoshimoto, M., Tajima, A., Nagayama, O., Yamaguchi, K., Goda, A., Yamashita, T., Sagara, K., Itoh, H., & Aizawa, T. (2006). Critical level of cerebral oxygenation during exercise in patients with left ventricular dysfunction. Circulation Journal, 70(11), 1457–1461. https://doi.org/10.1253/circj.70.1457

43. Koike, A., Itoh, H., Oohara, R., Hoshimoto, M., Tajima, A., Aizawa, T., & Fu, L. T. (2004b). Cerebral oxygenation during exercise in cardiac patients. Chest, 125(1), 182–190. https://doi.org/10.1378/chest.125.1.182

44. Labrecque, L., Drapeau, A., Rahimaly, K., Imhoff, S., Billaut, F., & Brassard, P. (2020). Comparable blood velocity changes in middle and posterior cerebral arteries during and following acute high-intensity exercise in young fit women. Physiological reports, 8(9), e14430. https://doi.org/10.14814/phy2.14430

45. Lassen, N. A. (1959). Cerebral blood flow and oxygen consumption in man. Physiological reviews, 39(2), 183–238. https://journals.physiology.org/doi/pdf/10.1152/physrev.1959.39.2.183

46. Lipnick, M. S., Cahill, E. A., Feiner, J. R., & Bickler, P. E. (2018). Comparison of transcranial doppler and ultrasound-tagged near infrared spectroscopy for measuring relative changes in cerebral blood flow in human subjects. Anesthesia & Analgesia, 126(2), 579–587. https://doi.org/10.1213/ane.0000000000002590

47. Lipton, P. (1999). Ischemic cell death in brain neurons. Physiological Reviews, 79(4), 1431–1568. https://doi.org/10.1152/physrev.1999.79.4.1431

48. Liu, T. T., & Brown, G. G. (2007). Measurement of cerebral perfusion with arterial spin labeling: Part 1. methods. Journal of the International Neuropsychological Society, 13, 1–9. https://doi.org/10.10170S1355617707070646

49. Logothetis, N. K. (2008). What we can do and what we cannot do with fMRI. Nature, 453(7197), 869–878. https://doi.org/10.1038/nature06976

50. Lucas, S. J. E., Ainslie, P. N., Murrell, C. J., Thomas, K. N., Franz, E. A., & Cotter, J. D. (2012). Effect of age on exercise-induced alterations in cognitive executive function: Relationship to cerebral perfusion. Experimental Gerontology, 47(8), 541–551. https://doi.org/10.1016/j.exger.2011.12.002

51. Lyngeraa, T. S., Pedersen, L. M., Mantoni, T., Belhage, B., Rasmussen, L. S., Lieshout, J. J., & Pott, F. C. (2013). Middle cerebral artery blood velocity during running. Scandinavian Journal of Medicine & Science in Sports, 23(1), e32–e37. https://doi.org/10.1111/sms.12009

52. MacIntosh, B. J., Shirzadi, Z., Scavone, A., Metcalfe, A. W. S., Islam, A. H., Korczak, D., & Goldstein, B. I. (2017). Increased cerebral blood flow among adolescents with bipolar disorder at rest is reduced following acute aerobic exercise. Journal of Affective Disorders, 208, 205–213. https://doi.org/10.1016/j.jad.2016.08.060

53. Malik, A. A., Williams, C. A., Weston, K. L., & Barker, A. R. (2018). Perceptual and prefrontal cortex haemodynamic responses to high-intensity interval exercise with decreasing and increasing work-intensity in adolescents. International Journal of Psychophysiology. https://doi.org/10.1016/j.ijpsycho.2018.07.473

54. Matsukawa, K., Ishii, K., Liang, N., Endo, K., Ohtani, R., Nakamoto, T., Wakasugi, R. Kadowaki, A., & Komine, H. (2015). Increased oxygenation of the cerebral prefrontal cortex prior to the onset of voluntary exercise in humans. Journal of Applied Physiology, 119(5), 452–462. https://doi.org/10.1152/japplphysiol.00406.2015

55. Moher, D., Shamseer, L., Clarke, M., Ghersi, D., Liberati, A., Petticrew, M., Shekelle, P., Stewart, L.A., PRISMA-P Group, 2015. Preferred reporting items for systematic review and meta-analysis protocols (PRISMA-P) 2015 statement. Systematic Reviews, 4(1), 1–9.

56. Monroe, D. C., Gist, N. H., Freese, E. C., O’Connor, P. J., McCully, K. K., & Dishman, R. K. (2016). Effects of sprint interval cycling on fatigue, energy, and cerebral oxygenation. Medicine & Science in Sports & Exercise, 48(4), 615–624. https://doi.org/10.1249/mss.0000000000000809

57. Moreau, D., & Chou, E. (2019). The acute effect of high-intensity exercise on executive function: A meta-analysis. Perspectives on Psychological Science, 14(5), 734–764. https://doi.org/10.1177/1745691619850568

58. Neary, J. P., Roberts, D. W., Leavins, N., Harrison, M. F., Croll, J. C., & Sexsmith, J. R. (2008). Prefrontal cortex oxygenation during incremental exercise in chronic fatigue syndrome. Clinical Physiology and Functional Imaging, 28(6), 364–372. https://doi.org/10.1111/j.1475-097x.2008.00822.x

59. Ogoh, S., Fadel, P. J., Zhang, R., Selmer, C., Jans, Ø., Secher, N. H., & Raven, P. B. (2005). Middle cerebral artery flow velocity and pulse pressure during dynamic exercise in humans. American journal of physiology: Heart and circulatory physiology, 288(4), H1526–H1531. https://doi.org/10.1152/ajpheart.00979.2004

60. Ogoh, S., & Ainslie, P. N. (2009). Regulatory mechanisms of cerebral blood flow during exercise: New concepts. Exercise and Sport Sciences Reviews, 37(3), 123–129. https://doi.org/10.1097/jes.0b013e3181aa64d7

61. Olivo, G., Nilsson, J., Garzón, B., Lebedev, A., Wåhlin, A., Tarassova, O., Ekblom, M., & Lövdén, M. (2021). Immediate effects of a single session of physical exercise on cognition and cerebral blood flow: A randomized controlled study of older adults. NeuroImage, 225. https://doi.org/10.1016/j.neuroimage.2020.117500

62. Parfitt, R., Hensman, M. Y., & Lucas, S. J. E. (2017). Cerebral blood flow responses to aquatic treadmill exercise. Medicine & Science in Sports & Exercise, 49, 1305–1312. https://doi.org/10.1249/MSS.0000000000001230

63. Perdomo, S. J., Balzer, J. R., Jakicic, J. M., Kline, C. E., & Gibbs, B. B. (2019). Acute effects of aerobic exercise duration on blood pressure, pulse wave velocity and cerebral blood flow velocity in middle-aged adults. Sport Sciences for Health, 15(3), 647–658. https://doi.org/10.1007/s11332-019-00566-w

64. Perentis, P. A., Cherouveim, E. D., Malliou, V. J., Margaritelis, N. V., Chatzinikolaou, P. N., Koulouvaris, P., Tsolakis, C., Nikolaidis, M. G., Geladas, N. D., & Paschalis, V. (2021). The effects of high-intensity interval exercise on skeletal muscle and cerebral oxygenation during cycling and isokinetic concentric and eccentric exercise. Journal of Functional Morphology and Kinesiology, 6(3), 62. https://doi.org/10.3390/jfmk6030062

65. Pontifex, M. B., Gwizdala, K. L., Weng, T. B., Zhu, D. C., & Voss, M. W. (2018). Cerebral blood flow is not modulated following acute aerobic exercise in preadolescent children. International Journal of Psychophysiology, 134, 44–51. https://doi.org/10.1016/j.ijpsycho.2018.10.007

66. Pugh, C. J. A., Sprung, V. S., Ono, K., Spence, A. L., Thijssen, D. H. J., Carter, H. H., & Green, D. J. (2015). The effect of water immersion during exercise on cerebral blood flow. Medicine & Science in Sports & Exercise, 47(2), 299–306. https://doi.org/10.1249/MSS.0000000000000422

67. Purkayastha, S., &Sorond, F. (2013). Transcranial doppler ultrasound: Technique and application. Semin Neurol, 32(4), 411–420. https://doi.org/10.1055/s-0032-1331812

68. Rattray, B., Smale, B. A., Northey, J. M., Smee, D. J., & Versey, N. G. (2017). Middle cerebral artery blood flow velocity during a 4 km cycling time trial. European Journal of Applied Physiology, 117(6), 1241–1248. http://dx.doi.org/10.1007/s00421-017-3612-2

69. Richardson, W. S., Wilson, M. C., Nishikawa, J., & Hayward, R. S. A. (1995). The well-built clinical question: A key to evidence-based decisions. American College of Physicians, 123, 1–12. https://doi.org/10.7326/ACPJC-1995-123-3-A12

70. Robertson, A. D., Crane, D. E., Rajab, A. S., Swardfager, W., Marzolini, S., Shirzadi, Z., Middleton, L. E., & Macintosh, B. J. (2015). Exercise intensity modulates the change in cerebral blood flow following aerobic exercise in chronic stroke. Experimental Brain Research, 233(8), 2467–2475. http://dx.doi.org/10.1007/s00221-015-4317-6

71. Rupp, T., & Perrey, S. (2008). Prefrontal cortex oxygenation and neuromuscular responses to exhaustive exercise. European Journal of Applied Physiology, 102(2), 153–163. https://doi.org/10.1007/s00421-007-0568-7

72. Santos-Concejero, J., Billaut, F., Grobler, L., Oliván, J., Noakes, T. D., & Tucker, R. (2017). Brain oxygenation declines in elite Kenyan runners during a maximal interval training session. European Journal of Applied Physiology, 117(5), 1017–1024. http://dx.doi.org/10.1007/s00421-017-3590-4

73. Sato, K., Ogoh, S., Hirasawa, A., Oue, A., & Sadamoto, T. (2011). The distribution of blood flow in the carotid and vertebral arteries during dynamic exercise in humans. The Journal of Physiology, 589(11), 2847–2856. https://doi.org/10.1113/jphysiol.2010.204461

74. Satterthwaite, T. D., Shinohara, R. T., Wolf, D. H., Hopson, R. D., Elliott, M. A., Vandekar, S. N., Ruparel, K., Calkins, M. E., Roalf, D. R., Gennatas, E. D., Jackson, C., Erus, G., Prabhakaran, K., Davatzikos, C., Detre, J. A., Hakonarson, H., Gur, R. C., & Gur, R. E. (2014). Impact of puberty on the evolution of cerebral perfusion during adolescence. Proceedings of the National Academy of Sciences, 111(23), 8643–8648. https://doi.org/10.1073/pnas.1400178111

75. Shibuya, K., Tanaka, J., Kuboyama, N., & Ogaki, T. (2004). Cerebral oxygenation during intermittent supramaximal exercise. Respiratory Physiology & Neurobiology, 140(2), 165–172. https://doi.org/10.1016/j.resp.2003.11.004

76. Simonson, S. G., & Piantidosi, C. A. (1996). Near-infrared spectroscopy: Clinical applications. Critical Care Clinics, 12(4), 1019–1029. https://doi.org/10.1016/s0749-0704(05)70290-6

77. Smirl, J. D., Hoffman, K., Tzeng, Y. C., Hansen, A., & Ainslie, P. N. (2016). Relationship between blood pressure and cerebral blood flow during supine cycling: Influence of aging. Journal of applied physiology, 120(5), 552–563. https://doi.org/10.1152/japplphysiol.00667.2015

78. Smith, B. A., Clayton, E. W., & Robertson, D. (2011). Experimental arrest of cerebral blood flow in human subjects: The red wing studies revisited. Perspectives in Biology and Medicine, 54(2), 121–131. https://doi.org/10.1353/pbm.2011.0018

79. Smith, K. J., & Ainslie, P. N. (2017). Regulation of cerebral blood flow and metabolism during exercise. Experimental Physiology, 102(11), 1356–1371. https://doi.org/10.1113/ep086249

80. Smith, K. J., MacLeod, D., Willie, C. K., Lewis, N. C. S., Hoiland, R. L., Ikeda, K., Tymko1, M. M., Donnelly, J., Day, T. A., MacLeod, N., Lucas, J. E., & Ainslie, P. N. (2014). Influence of high altitude on cerebral blood flow and fuel utilization during exercise and recovery. The Journal of Physiology, 592(24), 5507–5527. https://doi.org/10.1113/jphysiol.2014.281212

81. Smith, K. J., Suarez, I. M., Scheer, A., Chasland, L. C., Thomas, H. J., Correia, M., Dembo, L. G., Naylor, L. H., Maiorana, A. J., & Green, D. J. (2019). Cerebral blood flow during exercise in heart failure: Effect of ventricular assist devices. Medicine & Science in Sports & Exercise, 51, 1372–1379. https://doi.org/10.1249/MSS.0000000000001904

82. Sterne, J., Savović, J., Page, M. J., Elbers, R. G., Blencowe, N. S., Boutron, I., Cates, C. J., Cheng, H. Y., Corbett, M. S., Eldridge, S. M., Emberson, J. R., Hernán, M. A., Hopewell, S., Hróbjartsson, A., Junqueira, D. R., Jüni, P., Kirkham, J. J., Lasserson, T., Li, T., McAleenan, A., Reeves, B. C., Shepperd, S., Shrier, I., Stewart, L. A., Tilling, K., White, I. R., Whiting, P. F., & Higgins, J. (2019). RoB 2: A revised tool for assessing risk of bias in randomised trials. BMJ (Clinical research ed.), 366, l4898. https://doi.org/10.1136/bmj.l4898

83. Steventon, J. J., Foster, C., Furby, H., Helme, D., Wise, R. G., & Murphy, K. (2019). Hippocampal blood flow is increased after 20 min of moderate-intensity exercise. Cerebral Cortex. https://doi.org/10.1093/cercor/bhz104

84. Steventon, J. J., Furby, H., Ralph, J., O’Callaghan, P., Rosser, A. E., Wise, R. G., Busse, M., & Murphy, K. (2020). Altered cerebrovascular response to acute exercise in patients with Huntington’s disease. Brain Communications, 2(1). https://doi.org/10.1093/braincomms/fcaa044

85. Tallon, C. M., Simair, R. G., Koziol, A. V., Ainslie, P. N., & McManus, A. M. (2019). Intracranial vascular responses to high-intensity interval exercise and moderate-intensity steady-state exercise in children. Pediatric exercise science, 31(3), 290–295. https://doi.org/10.1123/pes.2018-0234

86. Tari, B., Shirzad, M., Behboodpour, N., Belfry, G. R., & Heath, M. (2021). Exercise intensity-specific changes to cerebral blood velocity do not modulate a postexercise executive function benefit. Neuropsychologia, 161, 108018. https://doi.org/10.1016/j.neuropsychologia.2021.108018

87. Thomas, H. J., Rana, U., Marsh, C. E., Caddy, H. T., Kelsey, L. J., Smith, K. J., Green, D. J., & Doyle, B. J. (2020). Assessment of cerebrovascular responses to physiological stimuli in identical twins using multimodal imaging and computational fluid dynamics. Journal of Applied Physiology, 129(5), 1024–1032. https://doi.org/10.1152/japplphysiol.00348.2020

88. Timinkul, A., Kato, M., Omori, T., Deocaris, C. C., Ito, A., Kizuka, T., Sakairi, Y., Nishijima, T., Asada, T., & Soya, H. (2008). Enhancing effect of cerebral blood volume by mild exercise in healthy young men: A near-infrared spectroscopy study. Neuroscience Research, 61(3), 242–248. https://doi.org/10.1016/j.neures.2008.03.012

89. Tontisirin, N., Muangman, S. L., Suz, P., Pihoker, C., Fisk, D., Moore, A., Lam, A. M., & Vavilala, M. S. (2007). Early childhood gender differences in anterior and posterior cerebral blood flow velocity and autoregulation. Pediatrics, 119(3), e610–e615. https://doi.org/10.1542/peds.2006-2110

90. Tsubaki, A., Morishita, S., Tokunaga, Y., Sato, D., Tamaki, H., Yamazaki, Y., Qin, W., & Onishi, H. (2018). Changes in cerebral oxyhaemoglobin levels during and after a single 20-minute bout of moderate-intensity cycling. Advances in Experimental Medicine and Biology, 1072. https://doi.org/10.1007/978-3-319-91287-5_20

91. Tsubaki, A., Takai, H., Oyanagi, K., Kojima, S., Tokunaga, Y., Miyaguchi, S., Sugawara, K., Sato, D., Tamaki, H., & Onishi, H. (2016). Correlation between the cerebral oxyhaemoglobin signal and physiological signals during cycling exercise: A near-infrared spectroscopy study. Advances in experimental medicine and biology, 923, 159–166. https://doi.org/10.1007/978-3-319-38810-6_21

92. Tsukamoto, H., Hashimoto, T., Olesen, N. D., Petersen, L. G., Sorensen, H., Nielsen, H.B., Secher, N. H., & Ogoh S. (2019). Dynamic cerebral autoregulation is maintained during high-intensity interval exercise. Medicine & Science in Sports & Exercise, 51, 372–378. https://doi.org/10.1249/MSS.0000000000001792

93. Van Lieshout, J. J., Wieling, W., Karemaker, J. M., & Secher, N. H. (2003). Syncope, cerebral perfusion, and oxygenation. Journal of Applied Physiology, 94(3), 833– 848. https://doi.org/10.1152/japplphysiol.00260.2002

94. Warpeha, J. (2003). Limitation of maximal oxygen consumption: The holy grail of exercise physiology or fool’s gold? Professionalization of Exercise Physiology online, 6(9), 1–16. https://www.asep.org/asep/asep/LimitationsMaximumOxygenConsumption.pdf

95. Williams, L. R., & Leggett, R. W. (1989). Reference values for resting blood flow to organs of man. Clinical Physics and Physiological Measurement, 10, 187–217. https://doi.org/10.1088/0143-0815/10/3/001

96. Willie, C. K., Tzeng, Y.-C., Fisher, J. A., & Ainslie, P. N. (2014). Integrative regulation of human brain blood flow. The Journal of Physiology, 592(5), 841–859. https://doi.org/10.1113/jphysiol.2013.268953

97. Witte, E., Liu, Y., Ward, J. L., Kempf, K. S., Whitaker, A., Vidoni, E. D., Craig, J., C., Poole, D. C., & Billinger, S. A. (2019). Exercise intensity and middle cerebral artery dynamics in humans. Respiratory Physiology & Neurobiology, 262, 32–39. https://doi.org/10.1016/j.resp.2019.01.013

